# Blood metabolites predicting Mild Cognitive Impairment in the Study of Latinos-Investigation of Neurocognitive Aging (HCHS/SOL)

**DOI:** 10.1101/2021.06.16.21258702

**Authors:** Shan He, Einat Granot-Hershkovitz, Ying Zhang, Jan Bressler, Wassim Tarraf, Bing Yu, Tianyi Huang, Donglin Zeng, Sylvia Wassertheil-Smoller, Melissa Lamar, Martha Daviglus, Maria J Marquine, Jianwen Cai, Thomas Mosley, Robert Kaplan, Eric Boerwinkle, Myriam Fornage, Charles DeCarli, Bruce Kristal, Hector M Gonzalez, Tamar Sofer

**Affiliations:** Department of Biostatistics, Harvard T.H Chan School of Public Health, Boston, MA, USA; Division of Sleep and Circadian Disorders, Brigham and Women’s Hospital, Boston, MA, USA; Department of Medicine, Harvard Medical School, Boston, MA, USA; Human Genetics Center, School of Public Health University of Texas Health Science Center at Houston, Houston, TX 77030, USA; Institute of Gerontology, Wayne State University, Detroit, MI, USA; Channing Division of Network Medicine, Brigham and Women’s Hospital, Boston, MA; Department of Biostatistics, Gillings School of Global Public Health, University of North Carolina, Chapel Hill, NC, USA; Department of Epidemiology & Population Health, Department of Pediatrics, Albert Einstein College of Medicine, Bronx, NY, USA; Department of Medicine, Institute for Minority Health Research, University of Illinois at Chicago, IL, USA; Rush Alzheimer’s Disease Research Center, Rush University Medical Center, Chicago, IL, USA; Department of Psychiatry, University of California, San Diego; Department of Medicine, University of Mississippi Medical Center, Jackson, Mississippi, USA; Division of Public Health Sciences, Fred Hutchinson Cancer Research Center, Seattle WA, USA; Human Genome Sequencing Center, Baylor College of Medicine, Houston, TX, USA; Brown Foundation Institute of Molecular Medicine, McGovern Medical School, The University of Texas Health Science Center at Houston, Houston, TX, USA; Alzheimer□s Disease Center, Department of Neurology, University of California, Davis, Sacramento, CA, USA; Burke Medical Research Institute, White Plains, New York, 10605, USA; Departments of Biochemistry and Neuroscience, Weill Medical College of Cornell University, New York, New York, 10021, USA; Department of Neurosciences and Shiley-Marcos Alzheimer’s Disease Center, University of California, San Diego, La Jolla, CA, USA

## Abstract

**INTRODUCTION:** Blood metabolomics-based biomarkers may be useful to predict measures of neurocognitive aging.

**METHODS:** We tested the association between 707 blood metabolites measured in 1,451 participants from the Hispanic Community Health Study/Study of Latinos (HCHS/SOL), with MCI and global cognitive change assessed seven years later. We further used Lasso penalized regression to construct a metabolomics risk score (MRS) that predicts MCI, potentially identifying a different set of metabolites than those discovered in individual-metabolite analysis.

**RESULTS:** We identified 20 metabolites associated with MCI and/or global cognitive change. Six of them were novel and 14 were previously reported as associated with neurocognitive aging outcomes. The MCI MRS comprised 61 metabolites and improved prediction accuracy from 84% (minimally adjusted model) to 89% in the entire dataset and from 75% to 87% among *APOE*-ε4 carriers.

**DISCUSSION:** Blood metabolites may serve as biomarkers identifying individuals at risk for MCI among U.S. Hispanics/Latinos.

## Introduction

Hispanics/Latinos in the U.S. are at higher risk for Mild cognitive impairment (MCI) and Alzheimer’s Disease and Related Dementias (ADRD), compared to non-Hispanic Whites, and are a rapidly growing ethnic population in the United States^1^. Various risk factors may explain the rate differences between populations, including genetic susceptibility, health conditions, lifestyle, and environmental factors^2^. For example, cardiovascular disease and diabetes are more prevalent in Hispanics/Latinos compared to non-Hispanic Whites and are known to increase the risk for ADRD. On the other hand, the strongest known genetic risk factor for ADRD, *APOE*-ε4, demonstrates a weaker association in Hispanics/Latinos, compared to other populations^3^.

Mild cognitive impairment (MCI) is a risk factor for the pathologic development of dementia^2^. Acceleration of the normal aging process of cognitive decline precedes the diagnosis of MCI^4^. Individuals diagnosed with MCI experience mild changes in thinking, memory, language, and/or judgment abilities greater than age-related expected changes. MCI can result from a variety of etiologies, some of which are reversible, and are affected by medications, injuries, sleep, exercise, education, and diet^5^. These MCI risk factors may lead to metabolic dysregulation. In the last decade, metabolome assessment has emerged as a new approach for biomarker discovery, and for evaluating the progress of disease and its underlying pathophysiology^6^. Recent studies have demonstrated risk prediction of MCI and ADRD based on blood metabolite biomarkers in prospective studies^7,8^. For example, higher plasma anthranilic acid levels were associated with a greater risk of dementia in the Framingham Offspring Study^9^. Thus, metabolome screening in middle-aged adults can detect plausible biomarkers that may improve risk prediction for MCI and can facilitate modifiable interventions at earlier stages of the disease^10 11^. However, most studies were conducted in non-Hispanic U.S. populations^7^, and metabolite associations may not generalize between populations. In one study, ten prospectively predictive phospholipids metabolites for MCI or dementia found in non-Hispanic Whites failed replication in an African American cohort^12^. Another study in European Americans^11^ replicated only one (out of seven) prospective predictive metabolite for dementia previously found in African Americans. Given the differences in risk factors and rates of cognitive decline and MCI across populations, it is plausible that optimal metabolomics biomarkers in Hispanics/Latinos are unique to this population.

We examined the associations of blood metabolites with MCI and global cognitive change over an average 7-years follow-up period among middle-aged and older adults in the Study of Latinos-Investigation of Neurocognitive Aging (SOL-INCA)^13^, an ancillary study to the Hispanic Community Health Study/Study of Latinos (HCHS/SOL). The analysis steps are described in Figure 1. We hypothesized that there are metabolites associated with MCI and cognitive decline in Hispanics/Latinos. We first tested the association of 707 metabolites with MCI and global cognitive change (Step A). We assessed the single-metabolite associations based on previous literature (Step B) and generalization to European and African Americans from the Atherosclerosis Risk In Communities (ARIC) study using a proxy-trait approach by testing metabolite associations with change in cognitive tests (Step C). Next, we constructed a metabolite risk score (MRS) using metabolites selected via a Lasso regression (Step D). Finally, we also explored whether previously reported metabolite associations with cognitive function generalize to the Hispanics/Latinos in the SOL-INCA target population (Step E).

**Figure 1:**
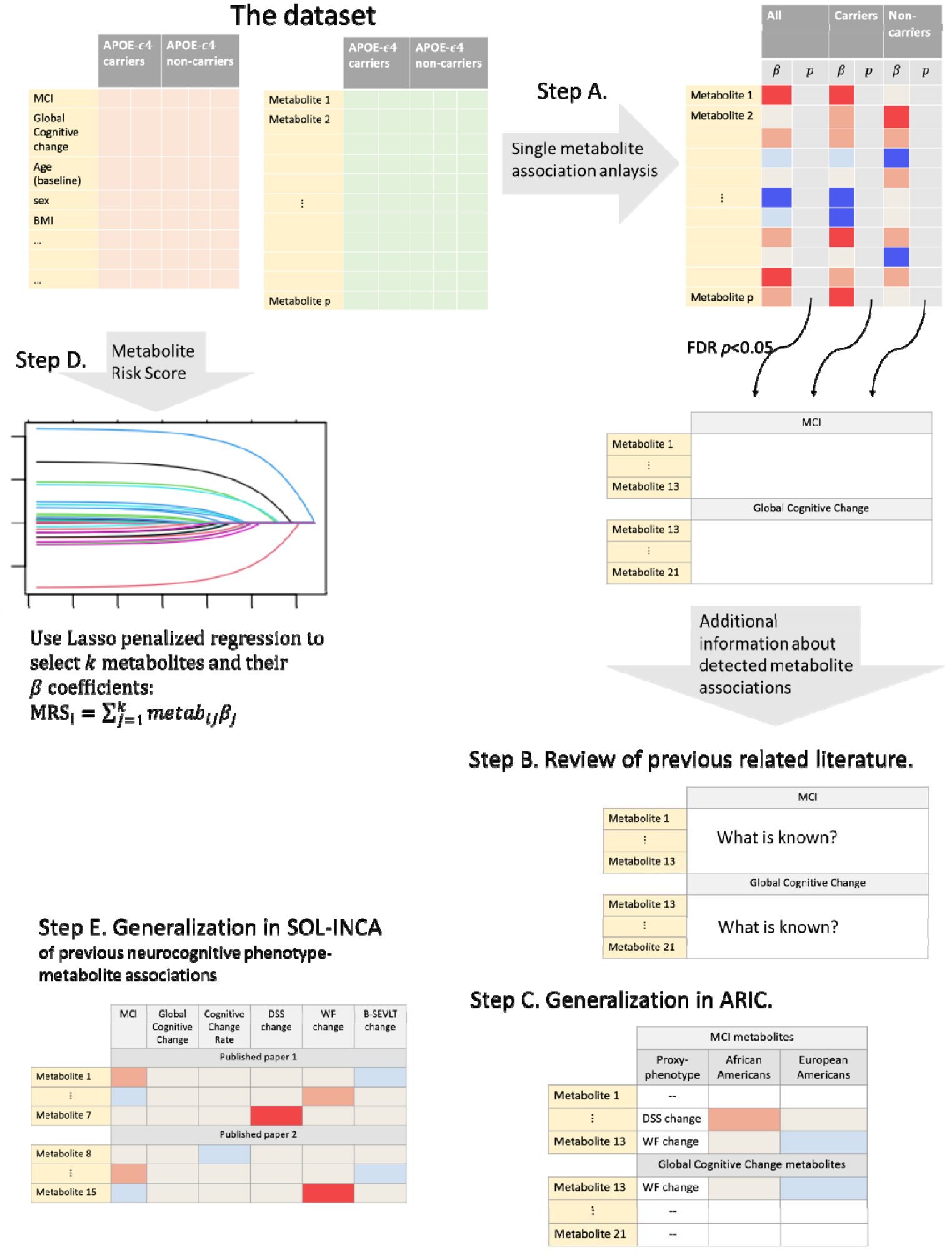
Metabolite - neurocognitive outcomes analyses flowchart in the SOL-INCA analytic dataset.

## Methods

### Study Population

The Hispanic Community Health Study/Study of Latinos (HCHS/SOL) is a U.S. population-based longitudinal cohort study with 16,415 Hispanic/Latino participants recruited from four field centers (Bronx, NY, Chicago, IL, Miami, FL, and San Diego, CA) by a sampling design previously described^14,15^. Adults ages 18-to 74-years-old were recruited during the first visit, between 2008 and 2011, and various biospecimen and health measures were collected. At baseline, cognitive function was assessed in 9,714 individuals aged 45 years or older. The baseline cognitive battery included the Six-Item Screener (SIS; mental status)^16^; Brief-Spanish English Verbal Learning Test (B-SEVLT; verbal episodic learning and memory)^17^; Word Fluency (WF; verbal functioning)^18^; and Digit Symbol Substitution test (DSS; processing speed, executive function)^19^. SOL-INCA is an ancillary study of HCHS/SOL, focusing on the middle-aged and older adult group who underwent cognitive assessment at Visit 1. Overall, 6,377 individuals 50-years or older with baseline cognitive testing participated in the SOL-INCA examination, taking place at or after HCHS/SOL visit 2, with an average of 7 years since Visit 1. Metabolites were measured in serum, after fasting, on a random subset of 3,978 from HCHS/SOL participants from visit 1. The current study population consists of 1,451 individuals who participated in SOL-INCA and additionally have measures of metabolites.

The details of the IRB/oversight body that provided approval or exemption for the research described are given below:

Mass General Brigham IRB

All necessary patient/participant consent has been obtained and the appropriate institutional forms have been archived.

### Neurocognitive outcomes

We studied two primary neurocognitive outcomes: prevalent Mild Cognitive Impairment (MCI) at the SOL-INCA visit, and global cognitive change in the 7-years follow-up between the HCHS/SOL Visit 1 and the SOL-INCA visits. Individuals were classified with MCI according to National Institute on Aging-Alzheimer’s Association criteria^20^. Details about the SOL-INCA MCI diagnostic operational procedures have been previously published^13,21^. The global cognitive change was computed using a confirmatory latent factor model using the HCHS/SOL baseline and SOL-INCA cognitive function tests, as previously described^13^. We performed secondary analyses in which we studied the association between the top identified metabolites and change in cognitive function measured by three cognitive tests corresponding to different domains: B-SEVLT for new learning and verbal memory, WF for verbal fluency, and DSS for psychomotor speed and sustained attention^20^. The changes were computed as the difference between the test scores in the SOL-INCA and visit 1. These associations were used for the analysis in Figure 1 Step C, as described later.

### Metabolomics measurement and processing

Metabolites were measured in serum of blood drawn after at least 8 hours of overnight fasting. Profiling was done using untargeted liquid chromatography-mass spectrometry (LC-MS) using the discovery HD4 platform in 2017 at Metabolon Inc. (Durham, NC). Of 1,136 metabolites quantified, 784 were identified as known compounds, and 352 were unidentified ^22^. Further details on metabolites’ quality control are provided in Supplementary Figure 1. Overall, we tested 707 metabolites in this study.

### Statistical analysis

We characterized the target population that the sample represents using weighted analysis, stratified by MCI. The weights are used to obtain estimates of characteristics generalizable to the target population of SOL-INCA. A detailed discussion of the sampling design, including the generation and use of weights for the HCHS/SOL was previously published^14,15^.

### Single metabolite association analyses

We tested the associations of each metabolite with each of the neurocognitive outcomes accounting for the complex survey design of the data using the R ‘survey’ package^23^, with a “quasi-Poisson” family for a binary outcome (Figure 1, Step A). We used three nested regression models, the first with basic adjustment for sex, age, study center, self-reported background (Dominican, Central American, Cuban, Mexican, Puerto Rican, South American, or more than one/other heritage), education, and years between HCHS/SOL visit 1 and the SOL-INCA visit. The second model with further adjustment for health measures including Body Mass Index (BMI), estimated Glomerular Filtration Rate (eGFR, estimated from serum creatinine using the CKD-EPI equation), type II diabetes (T2D), and hypertension. The third model with further adjustment for *APOE*-□4 carrier status (dominant mode), and lifestyle variables including alcohol consumption, and smoking status. We also stratified models 1 and 2 by *APOE*-□4 allele carrier status. The significance threshold was False Discovery Rate (FDR) adjusted p-value <0.05 (q-value) computed in each analysis separately. Biological pathways for metabolites were provided by Metabolon’s annotation file.

### Evidence from previous publications for single-metabolite identified associations

For each metabolite significantly associated with MCI or global cognitive change after multiple-testing correction, we searched for relevant published associations in Google Scholar, PubMed, PubChem, and The Human Metabolome Database (HMDB) using these keywords: the name/identification (e.g. HMDB ID) of the metabolite, “cognitive function”, “dementia”, “Alzheimer’s disease” (Figure 1 Step B).

### Generalization analysis of metabolite associations in the Atherosclerosis Risk in Communities (ARIC) study

We evaluated the generalizability of the discovered metabolite associations in the ARIC study (Figure 1, Step C), a longitudinal cohort study with cognitive measures and metabolite profiling based on a similar Metabolon platform. ARIC did not have equivalent measures of MCI and global cognitive change, therefore we used a proxy-generalization approach testing the metabolites’ associations with change in cognitive test results. Further details are provided in the Supplementary Materials.

### Metabolic Risk Score (MRS) for MCI

We performed Lasso-penalized regression to optimally select and estimate the joint effect of multiple metabolites that together predict MCI in the HCHS/SOL analytic sample (Step D in Figure 1)^24^. Based on the selected metabolites and their estimated effects, we constructed an MRS for MCI. Further details are provided in the Supplementary Materials.

### Generalization of previously reported metabolite associations with neurocognitive outcomes in SOL-INCA

We identified two manuscripts reporting associations of metabolites with MCI and AD, using a similar Metabolon platform^11,25^. To study whether their reported associations generalize to associations with MCI and global cognitive change in Hispanics/Latinos, we inspected the metabolites they reported in our single-metabolite association analyses (Figure 1, Step E).

### Data availability

HCHS/SOL and SOL-INCA data can be obtained from dbGaP under accession number phs000810.v1.p1. Summary statistics from association analyses of all metabolites and all studied phenotypes are available in GitHub repository. In addition, code for constructing the MCI MRS based on the Lasso-selected metabolites is provided in the same GitHub repository.

## Results

Table 1 characterizes demographic, health, and lifestyle characteristics of the study population, stratified by MCI status. Overall, our dataset included 1,451 individuals (558 men; 893 women), with a weighted mean age of 56 years at visit 1.

**Table 1:** Demographics, health, and lifestyle characteristics of the study population by MCI status.

|  | Without MCI | With MCI | Total |
| --- | --- | --- | --- |
| Sample Size | 1286 | 165 | 1451 |
| Age at visit 1 (mean (SD)) | 54.33 (7.05) | 58.12 (8.01) | 56.03 (8.18) |
| Gender = M (%) | 506 (49.1) | 52 (40.6) | 558 (48.1) |
| Study Center (%) |  |  |  |
| Bronx | 330 (30.0) | 40 (25.8) | 370 (29.5) |
| Chicago | 266 (10.1) | 38 (12.2) | 304 (10.3) |
| Miami | 386 (37.0) | 45 (41.4) | 431 (37.5) |
| San Diego | 304 (23.0) | 42 (20.6) | 346 (22.7) |
| Self-reported Background (%) |  |  |  |
| Dominican | 139 (11.6) | 16 (9.0) | 155 (11.3) |
| Central American | 134 (7.5) | 16 (9.0) | 150 (7.6) |
| Cuban | 239 (26.1) | 27 (27.4) | 266 (26.3) |
| Mexican | 461 (31.3) | 57 (24.8) | 518 (30.6) |
| Puerto Rican | 216 (16.0) | 35 (17.2) | 251 (16.1) |
| South American | 76 (4.4) | 9 (3.9) | 85 (4.3) |
| More than one/Other heritage | 20 (3.2) | 5 (8.7) | 25 (3.8) |
| Education Years (%) |  |  |  |
| <12 | 468 (32.7) | 85 (50.3) | 553 (34.8) |
| 12 | 294 (22.9) | 30 (12.9) | 324 (21.7) |
| >12 | 524 (44.4) | 50 (36.7) | 574 (43.4) |
| BMI (mean (SD)) | 29.91 (5.35) | 30.37 (5.50) | 29.50 (5.13) |
| eGFR (mean (SD)) | 90.64 (15.75) | 87.25 (15.96) | 88.72 (16.41) |
| Type II Diabetes (%) | 308 (25.4) | 68 (45.4) | 376 (27.8) |
| Hypertension (%) | 511 (42.0) | 91 (63.3) | 602 (44.5) |
| APOE-ε4 carriers (%) | 288 (22.0) | 41 (20.3) | 329 (21.8) |
| Alcohol consumption (n (%)) |  |  |  |
| Never | 249 (18.9) | 45 (27.9) | 294 (20.0) |
| Former | 403 (27.8) | 55 (28.6) | 458 (27.9) |
| Current | 634 (53.3) | 65 (43.5) | 699 (52.1) |
| Smoking status (n (%)) |  |  |  |
| Never | 726 (55.6) | 97 (51.1) | 823 (55.1) |
| Former | 324 (25.1) | 35 (24.5) | 359 (25.0) |
| Current | 236 (19.3) | 33 (24.4) | 269 (19.9) |
| Years between visits (mean (SD)) | 7.05 (1.16) | 7.17 (1.16) | 7.06 (1.16) |

Abbreviations: *MCI* mild cognitive impairment, *SD* standard deviation, *BMI* body mass index, *eGFR* Estimated glomerular filtration rate. (%) based on the sampling weights and complex survey design.
All measures are provided from visit 1.

### Single metabolite association analyses (Figure 1 Step A)

We performed metabolite association analysis across the total analytic sample and stratified by *APOE*-ε4 carrier status. Table 2 summarizes the associations that passed the FDR-adjusted threshold in any one of the three nested regression models. Across the three regression models, we identified 13 metabolites associated with MCI and 8 metabolites associated with global cognitive change. Effect directions persisted across the three models for all associations (data not shown). One metabolite, quinolinate, was associated with both MCI and global cognitive change.

**Table 2:** Single-metabolite associations with MCI or global cognitive decline in our analytic sample.

| Metabolites | Super pathway | All Participants |  |  |  | APOE-ε4 Carriers |  |  |  | APOE-ε4 Non-Carriers |  |  |  |
| --- | --- | --- | --- | --- | --- | --- | --- | --- | --- | --- | --- | --- | --- |
|  |  | Effect | SE | Unadj-p-value | FDR adj. p-value | Effect | SE | Unadj p-value | FDR adj. p-value | Effect | SE | Unadj. p-value | FDR adj. p-value |
| Outcome: MCI |  |  |  |  |  |  |  |  |  |  |  |  |  |
| MODEL 1 |  |  |  |  |  |  |  |  |  |  |  |  |  |
| 3-aminoisobutyrate | Nucleotide | -0.04 | 0.01 | 1.26E-02 | 1.00E+00 | -0.69 | 0.17 | 3.00E-05 | 1.76E-02 | -0.21 | 0.10 | 4.19E-02 | 1.00E+00 |
| 1-arachidonoyl-GPE (20:4n6) | Lipid | 0.03 | 0.01 | 1.44E-02 | 1.00E+00 | -0.20 | 0.15 | 1.80E-01 | 1.00E+00 | 0.40 | 0.10 | 4.00E-05 | 1.71E-02 |
| Glycolithocholic acid 3-sulfate | Lipid | 0.04 | 0.01 | 1.12E-03 | 1.00E+00 | -0.29 | 0.20 | 1.38E-01 | 1.00E+00 | 0.42 | 0.11 | 5.00E-05 | 1.71E-02 |
| 1-Palmitoyl-2-arachidonoyl-gpe (16:0/20:4) | Lipid | 0.03 | 0.01 | 6.63E-03 | 1.00E+00 | -0.04 | 0.16 | 7.82E-01 | 1.00E+00 | 0.40 | 0.11 | 2.20E-04 | 3.52E-02 |
| Quinolate | Cofactors and Vitamins | -0.03 | 0.01 | 6.66E-04 | 5.99E-01 | -0.13 | 0.17 | 4.37E-01 | 1.00E+00 | -0.33 | 0.09 | 4.30E-04 | 4.62E-02 |
| 9,10-DiHOME | Lipid | -0.04 | 0.01 | 4.16E-04 | 3.74E-01 | -0.13 | 0.20 | 5.23E-01 | 1.00E+00 | -0.38 | 0.11 | 5.10E-04 | 4.66E-02 |
| Glycodeoxycholatesulfate | Lipid | 0.04 | 0.01 | 8.64E-04 | 7.77E-01 | 0.02 | 0.20 | 9.23E-01 | 1.00E+00 | 0.34 | 0.09 | 1.70E-04 | 3.52E-02 |
| 2-hydroxyoctanoate | Lipid | -0.04 | 0.01 | 2.06E-04 | 1.86E-01 | -0.08 | 0.14 | 5.64E-01 | 1.00E+00 | -0.36 | 0.10 | 3.10E-04 | 3.93E-02 |
| MODEL 2 |  |  |  |  |  |  |  |  |  |  |  |  |  |
| 3-aminoisobutyrate | Nucleotide | -0.04 | 0.02 | 1.93E-02 | 1.00E+00 | -0.66 | 0.14 | 1.90E-01 | 8.50E-04 | -0.17 | 0.11 | 1.11E-01 | 1.00E+00 |
| 3-hydroxy-3-methylglutarate | Lipid | -0.02 | 0.01 | 9.42E-02 | 1.00E+00 | -0.68 | 0.16 | 1.00E-05 | 4.27E-03 | -0.06 | 0.11 | 6.16E-01 | 1.00E+00 |
| 1-arachidonoyl-GPE (20:4n6) | Lipid | 0.03 | 0.01 | 3.92E-02 | 1.00E+00 | -0.19 | 0.16 | 2.16E-01 | 1.00E+00 | 0.42 | 0.10 | 4.00E-05 | 2.69E-02 |
| Glycolithocholic acid 3-sulfate | Lipid | 0.04 | 0.01 | 2.60E-03 | 1.00E+00 | -0.27 | 0.18 | 1.46E-01 | 1.00E+00 | 0.39 | 0.11 | 2.20E-04 | 4.78E-02 |
| 1-palmitoleoyl-GPC (16:1) | Lipid | 0.03 | 0.02 | 4.40E-02 | 1.00E+00 | -0.20 | 0.13 | 1.14E-01 | 1.00E+00 | 0.42 | 0.11 | 1.60E-04 | 4.78E-02 |
| MODEL 3 |  |  |  |  |  |  |  |  |  |  |  |  |  |
| 3-aminoisobutyrate | Nucleotide | -0.04 | 0.02 | 1.38E-02 | 1.00E+00 | -0.68 | 0.16 | 1.00E-05 | 4.45E-03 | -0.19 | 0.11 | 1.00E-01 | 1.00E+00 |
| 3-hydroxy-3-methylglutarate | Lipid | -0.02 | 0.01 | 9.53E-02 | 1.00E+00 | -0.73 | 0.17 | 1.00E-05 | 4.45E-03 | -0.05 | 0.11 | 6.78E-01 | 1.00E+00 |
| Isocitrate | Energy | 0.01 | 0.01 | 5.44E-01 | 1.00E+00 | 0.57 | 0.16 | 2.60E-04 | 3.38E-02 | 0.04 | 0.12 | 7.49E-01 | 1.00E+00 |
| Sulfate | Xenobiotics | -0.01 | 0.01 | 4.19E-01 | 1.00E+00 | -0.52 | 0.14 | 1.70E-04 | 2.72E-02 | 0.08 | 0.12 | 4.82E-01 | 1.00E+00 |
| Tartarate | Xenobiotics | 0.01 | 0.01 | 7.72E-01 | 1.00E+00 | 0.52 | 0.14 | 1.70E-04 | 2.72E-02 | -0.14 | 0.10 | 1.64E-01 | 1.00E+00 |
| 1-arachidonoyl-GPE (20:4n6) | Lipid | 0.03 | 0.01 | 4.29E-02 | 1.00E+00 | -0.22 | 0.16 | 1.65E-01 | 1.00E+00 | 0.42 | 0.10 | 5.00E-05 | 2.01E-02 |
| Glycolithocholatesulfate | Lipid | 0.04 | 0.01 | 3.01E-03 | 1.00E+00 | -0.29 | 0.18 | 1.09E-01 | 1.00E+00 | 0.39 | 0.10 | 1.50E-04 | 3.15E-02 |
| 1-palmitoleoyl-GPC (16:1) | Lipid | 0.03 | 0.02 | 4.29E-02 | 1.00E+00 | -0.18 | 0.13 | 1.63E-01 | 1.00E+00 | 0.42 | 0.10 | 6.00E-05 | 2.01E-02 |

| Outcome: Global Cognitive Change |  |  |  |  |  |  |  |  |  |  |  |  |  |
| --- | --- | --- | --- | --- | --- | --- | --- | --- | --- | --- | --- | --- | --- |
| MODEL 1 |  |  |  |  |  |  |  |  |  |  |  |  |  |
| Kynurenine | Amino Acid | 0.14 | 0.03 | 4.00E-05 | 2.43E-02 | 0.04 | 0.06 | 5.24E-01 | 1.00E+00 | 0.16 | 0.04 | 6.00E-05 | 1.22E-02 |
| 7-Methylguanine | Nucleotide | 0.12 | 0.03 | 2.10E-04 | 4.45E-02 | 0.03 | 0.06 | 6.35E-01 | 1.00E+00 | 0.15 | 0.04 | 3.00E-05 | 1.22E-02 |
| Quinolate | Cofactors and Vitamins | 0.13 | 0.03 | 8.00E-05 | 2.43E-02 | 0.06 | 0.06 | 3.17E-01 | 1.00E+00 | 0.15 | 0.04 | 1.20E-04 | 1.86E-02 |
| Creatinine | Amino Acid | 0.12 | 0.04 | 3.46E-03 | 1.00E+00 | 0.02 | 0.07 | 7.99E-01 | 1.00E+00 | 0.16 | 0.05 | 4.20E-04 | 4.45E-02 |
| Gamma-glutamylleucine | Peptide | 0.11 | 0.03 | 6.60E-04 | 5.93E-01 | -0.02 | 0.07 | 7.51E-01 | 1.00E+00 | 0.16 | 0.04 | 4.00E-05 | 1.22E-02 |
| Gamma-glutamylvaline | Peptide | 0.13 | 0.04 | 3.53E-04 | 3.17E-01 | 0.06 | 0.06 | 3.71E-01 | 1.00E+00 | 0.15 | 0.04 | 4.90E-04 | 4.45E-02 |
| N-acetylcarnosine | Peptide | 0.16 | 0.05 | 4.95E-04 | 4.45E-01 | 0.09 | 0.08 | 2.60E-01 | 1.00E+00 | 0.19 | 0.05 | 2.90E-04 | 3.68E-02 |
| Thymol sulfate | Xenobiotics | 0.08 | 0.03 | 6.06E-03 | 1.00E+00 | -0.06 | 0.06 | 3.18E-01 | 1.00E+00 | 0.11 | 0.03 | 5.90E-04 | 4.72E-02 |

Abbreviations: *MCI* mild cognitive impairment, *SE* standard error. The positive effect for MCI is risk increasing, and the positive effect for global cognitive change is protective. MCI effect is log(odds ratio). Model 1: Adjusted for sex, age, study center, self-reported background, education, and duration in years between the visits. Model 2: Model 1 with further adjustment for BMI, eGFR, T2D, and Hypertension. Model 3: Model 2 with further adjustment for *APOE-ε4* carriers, alcohol consumption and smoking status.

Results from complete-cases metabolite analyses were similar to the analyses of the imputed metabolites. All but three associations were identified in one of the *APOE*-ε4 stratification groups (carriers or non-carriers), with most lipid metabolites identified in the *APOE*-ε4 non-carrier group. Figure 2 visualizes the correlations between the metabolites associated with MCI and/or global cognitive change as outlined in Table 2.

**Figure 2:**
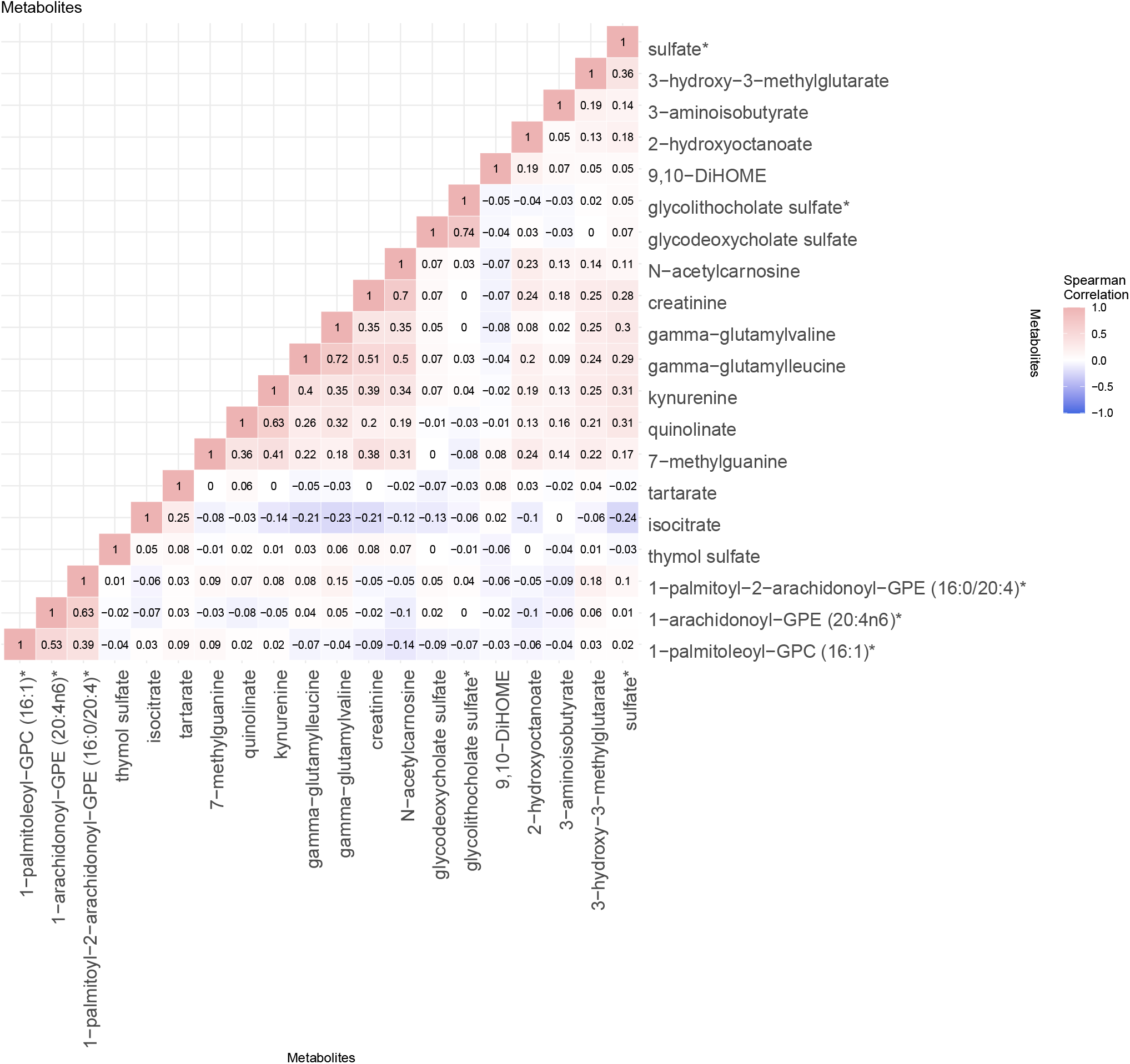
Correlations between significant FDR-adjusted metabolites associated with MCI or global cognitive decline. The metabolite correlations were computed within the total study population (not stratified by *APOE*-□4 carrier status).

Table 3 summarizes previously reported metabolite associations for the metabolites from Table 2 (Figure 1, Step B). Six metabolites associated with MCI in our study had no previously reported associations with neurocognitive phenotypes. Of these, three lipids were associated with MCI in *APOE*-ε4 non-carriers: 1-arachidonoyl-GPE (20:4n6), 2-hydroxyoctanoate, and 1-Palmitoyl-2-arachidonoyl-gpe (16:0/20:4). Two xenobiotics, tartarate and sulfate, and one nucleotide, aminoisobutyrate, were associated with MCI in *APOE*-ε4 carriers. All other 14 metabolites were previously reported as associated with relevant neurocognitive phenotypes in primarily non-Latino White and Black populations, 8 with a consistent direction of effects with the published literature, and 6 with opposite direction effects.

**Table 3:**
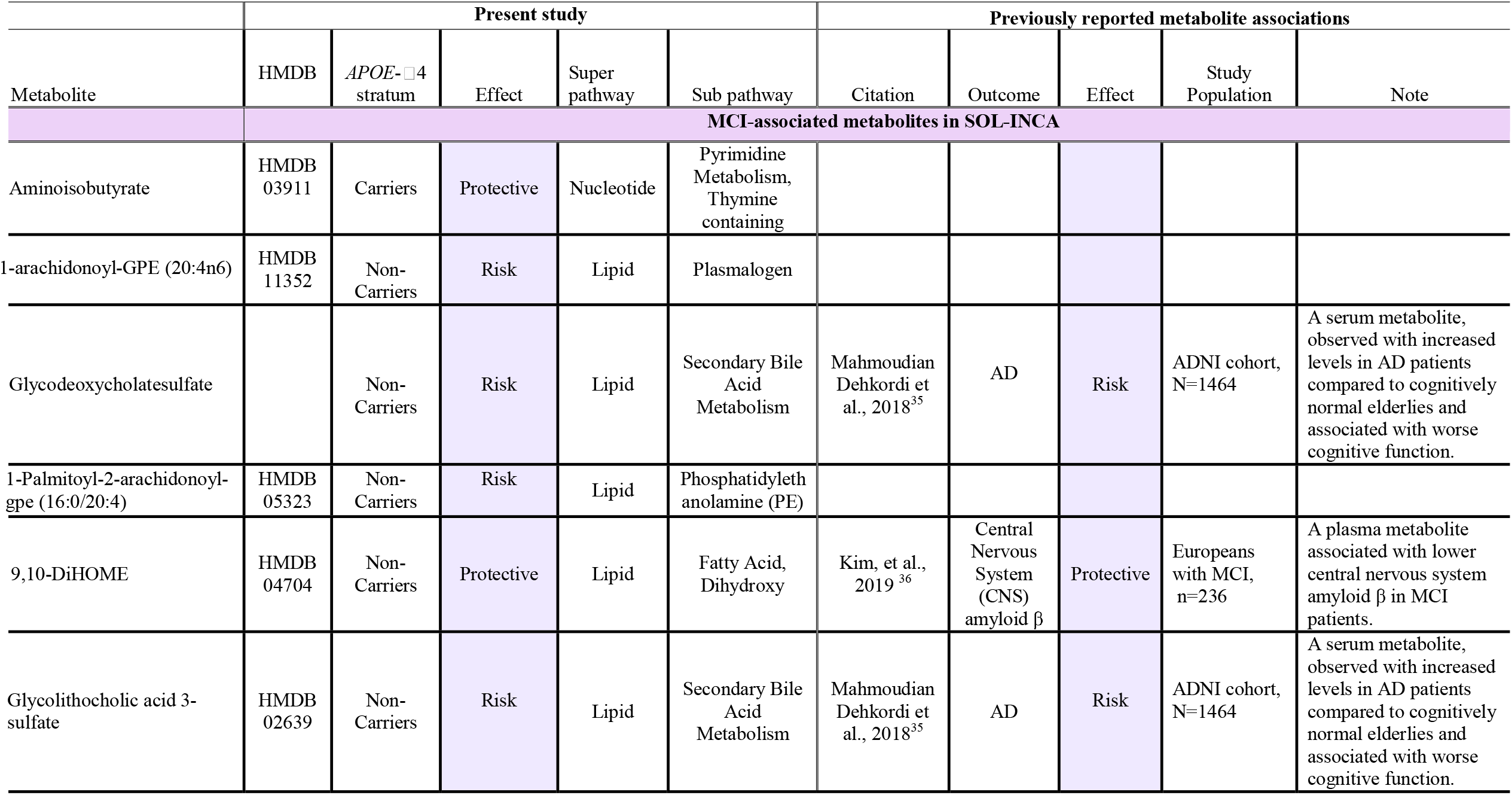
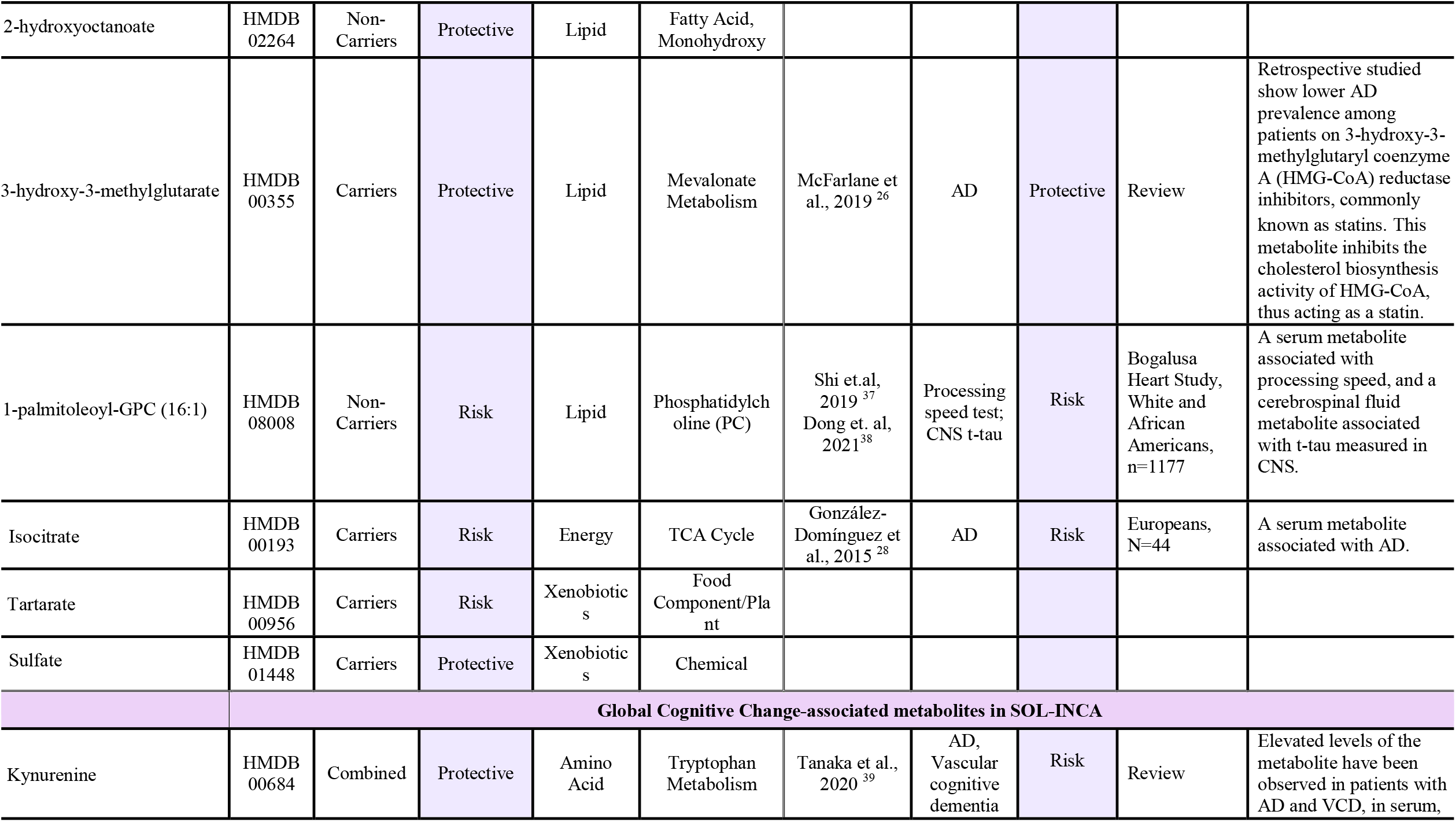
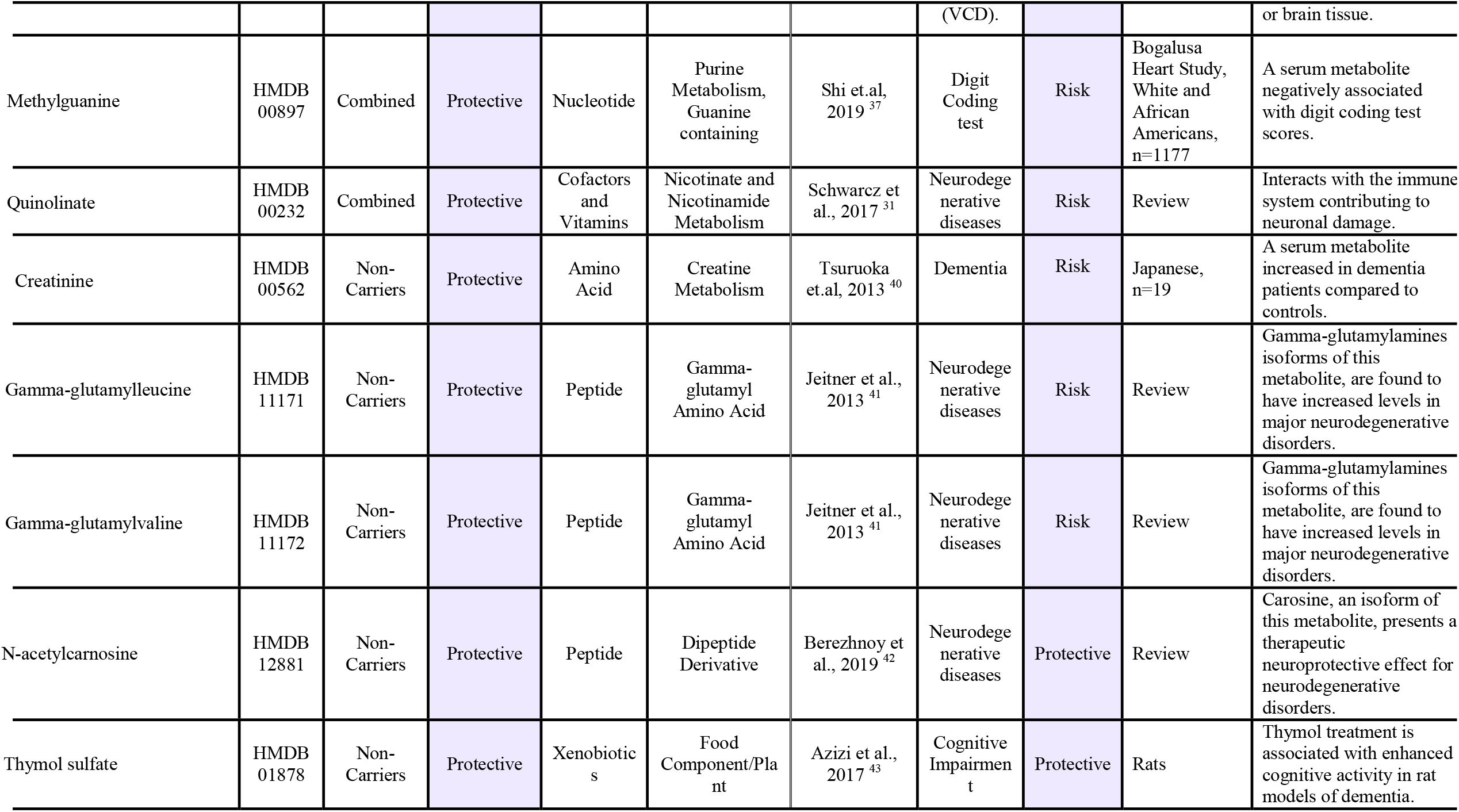
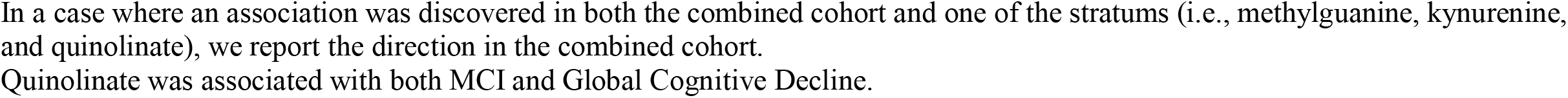
Previously reported metabolite associations for identified HCHS/SOL metabolite associations with cognitive functions.

### Generalization analysis of metabolite associations in ARIC (Figure 1, Step C)

Supplementary Tables 2-5 summarize proxy-phenotype selection for the ARIC cohort generalization study as described in the Supplementary Materials. Out of the 20 associated metabolites with either MCI or global cognitive change in our analytic dataset, 11 were available for ARIC generalization tests. Supplementary Table 6 summarizes the results from the generalization study in ARIC stratified by race: European American and African American. One association was generalized; 7-methylguanine was associated with improved cognitive change in the total analytic sample was associated with improved DSS change in African American ARIC *APOE*-ε4 carriers and non-carriers.

### Metabolic Risk Score (MRS) for MCI (Figure 1 Step D)

We performed Lasso regression to jointly select metabolites associated with MCI and estimate their effects. Sixty-one metabolites were selected by the algorithm and formed an MRS. Figure 3 compares MCI classification accuracy in a model accounting for baseline covariates only (sex, age, study center, self-reported Hispanic/Latino background, education, and time between visits), and a model further accounting for the MRS. Comparison of both models shows an increase in accuracy of 5% in the model which accounts for MRS within the total dataset, reaching an 89% accuracy. Within the *APOE*-ε4 carriers, the increased accuracy due to the addition of the MRS to the model is 12%, reaching 87% accuracy, and in the *APOE*-ε4 non-carriers, the increased accuracy is 7%, reaching a total of 90% accuracy. The MRS had a p-value < 0.0001 in all three models.

**Figure 3:**
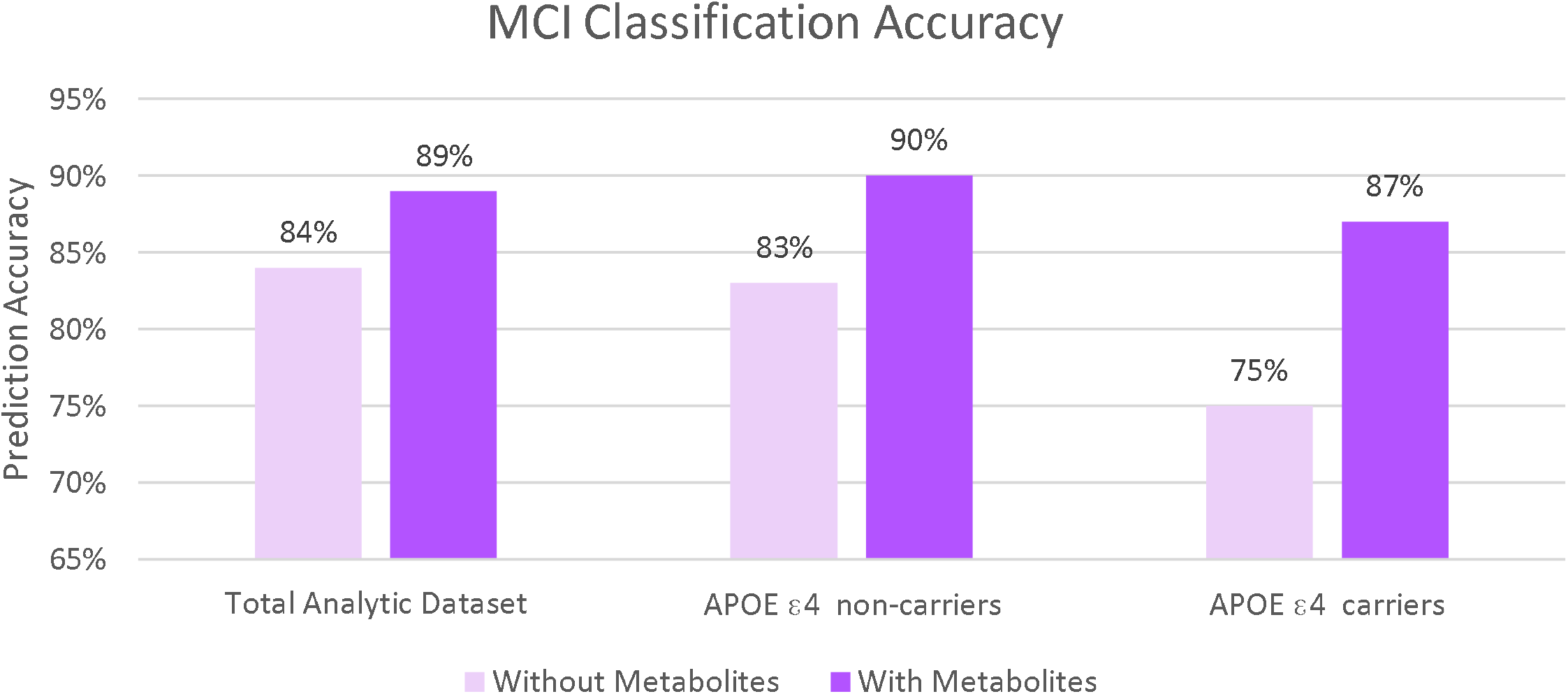
MCI Prediction accuracy for the total analytic dataset, and stratified by *APOE*-□4 status, for models adjusted for sex, age, study center, self-reported Hispanic/Latino background, education, and time between visits, with and without the metabolite risk score.

Of the 61 metabolites forming the MRS, four metabolites (aminoisobutyrate, quinolinate, 2-hydroxyoctanoate, and 1-palmitoleoyl-GPC (16:1)) were associated with MCI in single-metabolite analysis.

### Generalization of previously reported metabolite associations with neurocognitive outcomes to SOL-INCA Hispanics/Latinos (Figure 1 Step E)

Supplementary Figure 3 provides generalization results of 14 previously reported associations of Metabolon (Durham, NC, USA) measured metabolites available for SOL-INCA, with all primary and secondary neurocognitive outcomes. The metabolites were reported either in the African-American population or in a primarily White population. Five associations were generalized (one-sided p-value < 0.05) to MCI: androstenediol, docosapentaenoate, glycodeoxycholate, taurodeoxycholate were associated with increased risk of MCI (previously associated increased risk of dementia, AD, and reduced performance in DSS over time), and ursodeoxycholate was protective against MCI (previously reported as protective against AD).

## Discussion

We studied the association of 707 serum metabolites with MCI and global cognitive change phenotypes in Hispanics/Latinos from SOL-INCA. Overall, we identified 20 metabolite associations with either one or both of the phenotypes, mostly detected in *APOE*-ε4 non-carriers. To the best of our knowledge, 6 of the metabolites are novel associations and 14 had previously published evidence of association with neurocognitive phenotypes, from either diverse population-based, clinical, animal, or in-vitro studies. We constructed an MRS based on 61 metabolites to form a single score predictive of MCI, improving the accuracy of minimally-adjusted MCI prediction models in our Hispanics/Latinos. Finally, we generalized to our Hispanics/Latinos 5 out of 14 metabolite associations with neurocognitive phenotypes reported in previous publications that used the same metabolomics platform. All generalized associations were of lipid metabolites.

There were 13 metabolites associated with MCI after FDR control in our analysis. Of these, eight were lipids, and seven of them were identified in *APOE-*ε4 non-carriers (Table 2). As outlined in Table 3, five of the eight lipids were previously reported as associated with neurocognitive phenotypes with similar directions of associations (glycolithocholic acid 3-sulfate, glycodeoxycholatesulfate, 1-palmitoleoyl-GPC (16:1), 3-hydroxy-3-methylglutarate, and 9,10-DiHOME). The single lipid association detected in *APOE-*ε4 carriers was with 3-hydroxy-3-methylglutarate previously reported as a protective factor for AD26. Lipids are known for their crucial function in cell signaling, physiological processes, and disease pathology, especially in the brain^27^. *APOE*-ε4 genotype is known to disrupt lipid transport and metabolism in Alzheimer’s disease (AD). Our results from the *APOE*-ε4 non-carriers emphasize the importance of lipids dysregulation in neurocognitive phenotypes, even in the absence of the *APOE*-ε4 allele^27^. Isocitrate is another metabolite associated with increased MCI risk in *APOE*-ε4 carriers. It was previously reported as having a higher concentration in serum of AD patients compared to cognitively normal older adults ^28^. Isocitrate is a part of the citrate cycle pathway, responsible for the oxidation of carbohydrates and fatty acids.

We identified 8 metabolites associated with global cognitive change, all were previously implicated with neurocognitive phenotypes (Table 3). The directions of six of the associations were inconsistent with the literature, being protective in the present study whereas associated with increased risk in previous reports. All these six metabolites also were highly positively correlated in our dataset (Figure 2). The direction of association in our dataset remained consistent after adjusting for additional neurocognitive risk factors in regression models 2 and 3, though they became less statistically significant. One of these metabolites, 7-methylguanine was also associated with improved cognitive function (change in DSS test) in ARIC African Americans (p-values = 0.03 and 0.04 in *APOE*-ε4 carriers and non-carriers, respectively). The inconsistencies between our findings and prior literature could result from different neurocognitive phenotypes evaluated across studies, differences in population characteristics such as age and genetic architecture, or other lifestyle and environmental factors that could not be adequately accounted for in the models^29^. Such differences between populations may also explain the overall low generalization in the ARIC population (Supplementary Table 6). Further investigation of these metabolites in additional data collected in Hispanics/Latinos, and across diverse populations, is needed to clarify the role of these metabolites in neurocognitive health.

A single metabolite, quinolinate, was associated with both MCI and cognitive change in our analysis. Quinolinate is part of the kynurenine pathway of tryptophan catabolism, involved in response to inflammation and infection. The kynurenine pathway is increasingly recognized as contributing to cognitive function^30,31^. Higher concentrations of both quinolinate and kynurenine were associated with improved cognitive change in our study, while previous studies primarily reported these two metabolites as associated with neuronal damage31. In generalization analysis in European Americans from ARIC, quinolinate was nominally associated (p-value = 0.05) with worse 6-year cognitive performance in the DSS test in *APOE*-ε4 carriers. A possible explanation for these contrasting results could be the recognition that quinolinate is a double-edged sword, acting as both an essential metabolite in the kynurenine pathway of tryptophan catabolism and a potent neurotoxin with a pro-apoptotic effect on some cell types^32^.

We identified 6 novel associations of metabolites with MCI that were not previously implicated with neurocognition. Aminoisobutyrate, a protective factor, is a nucleotide in the pyrimidine metabolism pathway produced by skeletal muscle during physical activity. This metabolite was suggested to protect from metabolic syndrome and its cardiovascular complications, which are known risk factors for dementia^33^. Three other metabolites are lipids: risk-increasing 1-arachidonoyl-GPE (20:4n6), protective 2-hydroxyoctanoate, risk-increasing 1-Palmitoyl-2-arachidonoyl-gpe (16:0/20:4), and the two other metabolites are xenobiotics, chemicals that are not derived in humans and enter the body via food or other environmental exposure, risk-increasing tartarate and protective sulfate. Of these, only the sulfate was available for generalization analysis in ARIC, with DSS as proxy-phenotype, and there was no evidence of association (p-value >0.5 in both European and African Americans). Tartarate is an organic acid occurring in many fruits, thus it is regulated in the body by dietary consumption. Similarly, the quinolinate mentioned above is regulated by dietary consumption of vitamin B3^34^, thus supporting the investigation of diet as a risk factor for neurocognitive outcomes.

The MCI MRS created by Lasso comprised 61 metabolites, and MCI prediction models that included the MRS improved prediction accuracy compared to the minimally adjusted models for both the entire dataset and among the *APOE*-ε4 carriers and non-carriers. Future targeted data collection is needed to test the performance of the MRS in the prediction of MCI in an independent dataset with similar and/or different ancestries. Validation using existing datasets is challenging because measured metabolites often do not overlap between different studies, in addition to differences in measured phenotypes. Since MCI is often a transitional stage before the development of dementia, the MRS, if validated, could potentially be used as an early detection biomarker allowing for prevention strategies.

The present study adds to the growing body of research that utilizes fasting blood metabolites to predict neurocognitive aging outcomes. We provide an in-depth analysis of the associations of a broad panel of metabolites with neurocognitive outcomes studied in a unique prospective cohort comprised solely of the U.S. Hispanic/Latino understudied population. However, our study also has some limitations. First, despite the relatively large HCHS/SOL dataset, comprising 1451 participants, only 10% of them were classified as MCI, due to their relatively young age, thus limiting statistical power. Moreover, in association analysis within the stratum of *APOE*-□4 carriers, the sample size is quite small (n=329), limiting the power and potential generalizability to other populations, and increasing the likelihood of overfitting. Second, despite accounting for several covariates in the nested models, these covariates may not fully capture the relationship of lifestyle and environmental confounders with both metabolites and cognitive outcomes, resulting in additional unobserved confounding. Finally, we were not able to fully test our novel metabolite in an independent study population. The ARIC generalization study was also conducted in young adults, used different phenotypes, and had only one of the six novel metabolites detected in the present study. Nevertheless, most of our findings validated previous metabolite associations with cognitive functions, offering external validity. Further analyses with other Hispanic/Latino data are needed to replicate our novel associations.

Overall, this study shows evidence that optimal metabolomics biomarkers for MCI and global cognitive change in Hispanics/Latinos are different than those in Whites and other populations. We found novel associated metabolites that may be specific to our population and replicated previously published metabolite associations, but some showed the opposite direction of associations. Further work should study the similarity and differences in metabolomics biomarkers predicting neurocognitive phenotypes across diverse populations.

## Data Availability

HCHS/SOL and SOL-INCA data can be obtained from dbGaP under accession number phs000810.v1.p1. Summary statistics from association analyses will become available upon publication of the peer-reviewed manuscript.

## Acknowledgments

The Hispanic Community Health Study/Study of Latinos is a collaborative study supported by contracts from the National Heart, Lung, and Blood Institute (NHLBI) to the University of North Carolina (HHSN268201300001I / N01-HC-65233), University of Miami (HHSN268201300004I / N01-HC-65234), Albert Einstein College of Medicine (HHSN268201300002I / N01-HC-65235), University of Illinois at Chicago (HHSN268201300003I / N01-HC-65236 Northwestern Univ), and San Diego State University (HHSN268201300005I / N01-HC-65237). The following Institutes/Centers/Offices have contributed to the HCHS/SOL through a transfer of funds to the NHLBI: National Institute on Minority Health and Health Disparities, National Institute on Deafness and Other Communication Disorders, National Institute of Dental and Craniofacial Research, National Institute of Diabetes and Digestive and Kidney Diseases, National Institute of Neurological Disorders and Stroke, NIH Institution-Office of Dietary Supplements. This work was supported by the National Institute on Aging (R01AG048642, RF1AG054548, RF1AG061022, and R21AG056952). Dr. González also receives additional support from P30AG062429 and P30AG059299. Support for metabolomics data was graciously provided by the JLH Foundation (Houston, Texas). The Atherosclerosis Risk in Communities study has been funded in whole or in part with Federal funds from the National Heart, Lung, and Blood Institute, National Institutes of Health, Department of Health and Human Services (contract numbers HHSN268201700001I, HHSN268201700002I, HHSN268201700003I, HHSN268201700004I, and HHSN268201700005I). We thank the staff and participants of the ARIC study for their important contributions. The metabolomics research was sponsored by the National Human Genome Research Institute (3U01HG004402-02S1).

