## Supplemental data for "Blood metabolites predicting Mild Cognitive Impairment in the Study of Latinos-Investigation of Neurocognitive Aging (HCHS/SOL)"

Supplementary Materials

Shan He, Einat Granot-Hershkovitz, Ying Zhang, Jan Bressler, Wassim Tarraf, Bing Yu, Tianyi Huang, Donglin Zeng, Sylvia Wassertheil-Smoller, Melissa Lamar, Martha Daviglus, Maria J Marquine, Jianwen Cai, Thomas Mosley, Robert Kaplan, Eric Boerwinkle, Myriam Fornage, Charles DeCarli, Bruce Kristal, Hector M Gonzalez, Tamar Sofer

Metabolites (n=784)

Metabolites with <25% missing (n=642)

Complete-Case

Rank-Normalize

Impute: Half the minimum observed

Rank-Normalize

Metabolites with 25%-75% missing (n=67)

Dichotomized into observed vs non-observed

Metabolites with >75% missing (n=75)

Excluded from analysis

Supplementary Figure 1: Analytic approach to the metabolites measured in our study sample. As a quality control measure, we excluded metabolites with more than 75% missing values across the sample, and dichotomized metabolites with 25-75% missing values into “observed” and “not observed”. Metabolites with less than 25% missing were treated as continuous. For these, we performed two analyses: the primary discovery analyses used imputed metabolite values for individuals with missing values, and for a secondary analysis we used only complete cases. For a given metabolite, missing values were imputed using half of the lowest non-missing value observed for that metabolite in the analytic sample, under the assumption that values were missing due to abundance lower than the detection limit. Finally, metabolites that were treated as continuous variables were rank-normalized (with ties being replaced by their means) to reduce potential false detections resulting from high leverage values. In total, we tested 707 metabolites in this current study.

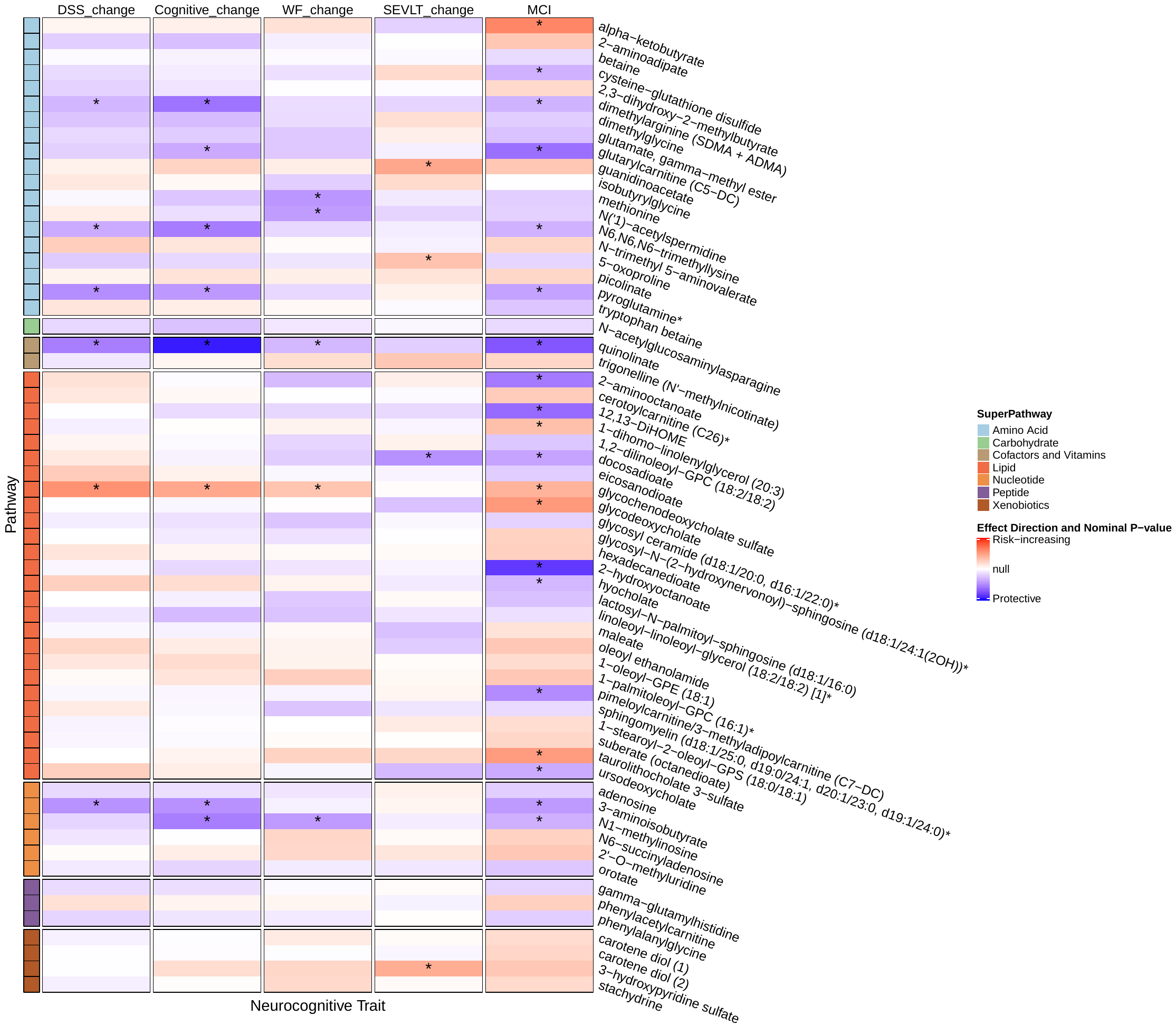

Supplementary Figure 2: Lasso Regression selected metabolites’ association with neurocognitive phenotypes: Mild Cognitive Impairment (MCI), Global Cognitive Decline, Digit Symbol Substitution Test (DSS), Brief-Spanish English Verbal Learning Test (SEVLT), and Word Fluency (WF). 61 metabolites were selected and aligned according to their corresponding super pathway: 1 - Amino Acid, 2 - Carbohydrate, 3 - Cofactors and Vitamins, 4 - Lipid, 5 - Nucleotide, 6 - Peptide, 7 - Xenobiotics. Asterisks represent nominal associations (p-value < 0.05) when testing the association of each metabolite individually against the cognitive Outcome. Heatmap colors correspond to directions of association. The strength of the colors corresponds to the strength of the evidence of association, quantified by -log(p-value), so that white color is obtained when the p-value is 1, and darker colors are obtained for lower p-values. The log(p-value) scale was selected, rather than effect sizes, due to different scales across the neurocognitive phenotypes.

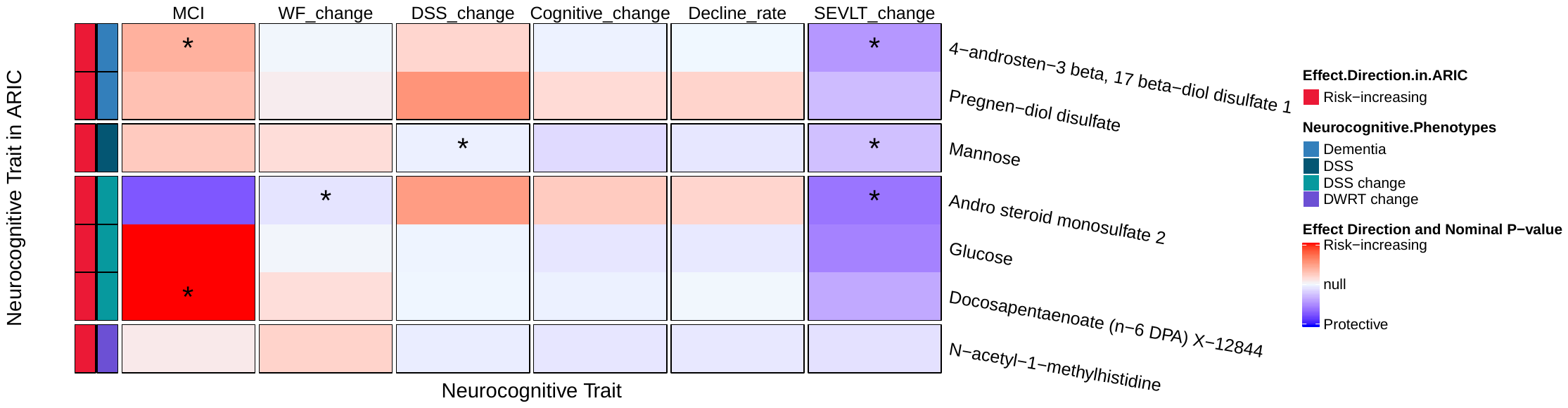

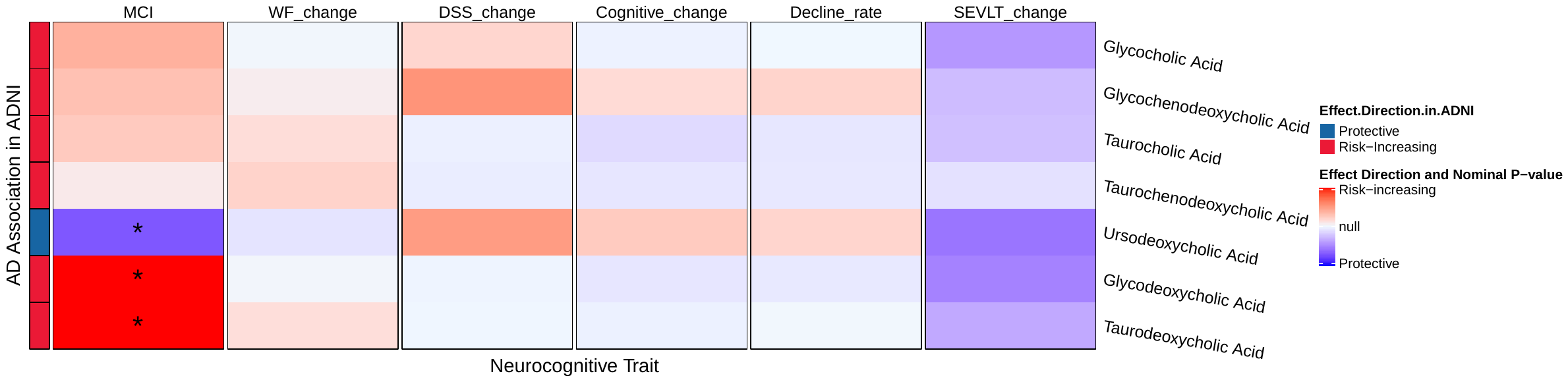

Abbreviations: MCI Mild Cognitive Impairment, DSS Digit Symbol Substitution Test, DWRT Delayed Word Recall Test, WF word fluency test score, SEVLT Spanish English Verbal Learning Test, ADNI Alzheimer's Disease Neuroimaging Initiative, AD Alzheimer’s Disease

* Nominally significant results.

Supplementary Figure 3: Generalization of previously reported metabolites association with neurocognitive relevant phenotypes to SOL-INCA Hispanics/Latinos.

### ARIC Methods

The Atherosclerosis Risk in Communities (ARIC) study is a prospective longitudinal study of the development of atherosclerosis, and its clinical sequelae in which 15,792 individuals aged 45-64 years from four communities in the United States were enrolled at the baseline examination (1987-1989). A detailed description of the ARIC study has been reported previously^1^. Various medical information and specimens were collected at each of the eight examinations. In particular, cognitive testing for this study was performed in the entire cohort at the 2^nd^ and 4^th^ examinations in 1990-1992 and 1996-1998, respectively. Written informed consent was provided by all study participants, and the study design and methods were approved by the institutional review boards at each of the collaborating medical institutions: University of Mississippi Medical Center Institutional Review Board (Jackson Field Center); Wake Forest University Health Sciences Institutional Review Board (Forsyth County Field Center); University of Minnesota Institutional Review Board (Minnesota Field Center); and Johns Hopkins University School of Public Health Institutional Review Board (Washington County Field Center). In this analysis, we use 1,151 European American and 325 African American ARIC participants who had measured metabolites in serum from their 1^st^ clinic visit. We excluded participants who had (1) unknown history of definite or probable stroke or a history of physician-diagnosed stroke prior to visit 2, or (2) had missing information on education at baseline. Detailed descriptions of participants' health, demographic, and lifestyle characteristics are presented below.

| **Characteristics** | **European American** | **African American** |
| --- | --- | --- |
| Sample Size | 1151 | 325 |
| Age at visit 2 (mean (SD)) | 57.28 (5.70) | 55.92 (5.31) |
| Gender = M (N, %) | 514 (44.7) | 114 (35.1) |
| Study Center (N, %) |  |  |
| Forsyth County, NC | 328 (28.5) | 83 (25.5) |
| Minneapolis, MN | 398 (34.6) | N/A |
| Washington County, MD | 425 (36.9) | N/A |
| Jackson, MS | N/A | 242 (74.5) |
| BMI (kg/m^2^) (mean (SD)) | 27.62 (5.30) | 30.13 (6.24) |
| Education (N, %) |  |  |
| <12 | 180 (15.6) | 116 (35.7) |
| 12 | 401 (34.9) | 70 (21.5) |
| >12 | 570 (49.5) | 139 (42.8) |
| Alcohol consumption (N, %) |  |  |
| Never | 209 (18.1) | 102 (31.4) |
| Former | 193 (16.8) | 105 (32.3) |
| Current | 749 (65.1) | 118 (36.3) |
| Smoking (N, %) |  |  |
| Never | 452 (39.3) | 145 (44.6) |
| Former | 479 (41.6) | 101 (31.1) |
| Current | 220 (19.1) | 79 (24.3) |
| eGFR (visit 1) (mean (SD)) | 91.93 (13.81) | 102.08 (17.68) |
| Hypertension (N, %) | 344 (29.9) | 163 (50.2) |
| Diabetes (N, %) | 107 (9.3) | 66 (20.3) |
| *APOE-ɛ4 (at least 1 allele) (N, %)* | 358 (31.1) | 133 (40.9) |
| Years between visits (mean (SD)) | 6.03 (0.28) | 6.04 (0.48) |

Abbreviations: *SD* standard deviation, *BMI* body mass index, *eGFR* Estimated glomerular filtration rate.

### Identifying proxy phenotypes for ARIC generalization

Since ARIC does not have equivalent MCI nor global cognitive decline outcomes, we performed proxy-phenotype generalization, in which we used the phenotypes that are available in the two cohorts, namely, change in B-SEVLT, WF, and DSS test, or equivalently in ARIC, change in Delayed Word Recall Test (DWRT), Word Fluency Test (WFT), and Digit Symbol Substitution Test (DSST), respectively^2^. Similarly, to the SOL-INCA methodology, a six-year change in cognitive function was computed as the difference between the test score obtained at visit 4 and the test score obtained at visit 2 for each test. The average change in DWRT was -0.2 (SD=1.52) for European Americans and -0.18 (SD=1.66) for African Americans. The average change in DSST was -2.79 (SD=6.03) for European Americans and -1.66 (SD=8.03) for African Americans. The average change in WFT was -0.52 (SD=7.8) for European Americans and -1 (SD=7.15) for African Americans.

For the significant associations highlighted in table 2 in the main manuscript, we searched for a proxy-phenotype of the change phenotypes above, as follows. For a significant metabolite-phenotype association detected in primary analysis:

1. Estimate the association of the metabolite with change phenotypes defined above in HCHS/SOL (B-SEVLT, WF, and DSS).
2. Identify associations with p-value < 0.05 (Supplement Tables 1-3). If none exists there is no proxy phenotype. Otherwise, change phenotypes with p-value < 0.05 are potential proxy phenotypes.
3. If there are several potential proxy-phenotype, choose the one with the lowest p-value in association with the metabolite. This is the selected proxy phenotype.

The proxy phenotypes for each of the associations in Table 2 are provided in Supplementary Table 4.

For the metabolite with proxy-phenotype matched, we used the same regression model in ARIC. Association was determined generalized if its nominal one-sided p-values guided by the direction of association in HCHS/SOL was less than 0.05.

We confirmed the compatibility of the blood serum metabolites names between the two studies, HSHC/SOL and ARIC according to HMDB ID, as summarized in the following table:

| MCI | | Global Cognitive Decline | |
| --- | --- | --- | --- |
| SOL-INCA | ARIC | SOL-INCA | ARIC |
| 3-aminoisobutyrate | NA | kynurenine | kynurenine |
| 1-arachidonoyl-GPE (20:4n6) * | 1-arachidonoylglycerophosphoethanolamine | 7-methylguanine | 7-methylguanine |
| glycolithocholate sulfate* | glycolithocholate sulfate | quinolinate | quinolinate |
| 1-palmitoyl-2-arachidonoyl-GPE (16:0/20:4) * | NA | creatinine | creatinine |
| quinolinate | quinolinate | gamma-glutamylleucine | gamma-glutamylleucine |
| 9,10-DiHOME | NA | gamma-glutamylvaline | gamma-glutamylvaline |
| glycodeoxycholate sulfate | NA | N-acetylcarnosine | N-acetylcarnosine |
| 2-hydroxyoctanoate | 2-hydroxyoctanoate | thymol sulfate | thymol sulfate |
| 3-hydroxy-3-methylglutarate | 3-hydroxy-3-methylglutarate |  |  |
| 1-palmitoleoyl-GPC (16:1) * | 1-palmitoleoylglycerophosphocholine (16:1) |  |  |
| isocitrate | NA |  |  |
| sulfate | sulfate |  |  |
| tartarate | tartarate |  |  |
| Proportion Matched | 8/13 | Proportion Matched | 8/8 |

Abbreviations: *MCI* mild cognitive impairment, *SOL-INCA* Study of Latinos-Investigation of Neurocognitive Aging, *ARIC* Atherosclerosis Risk In Communities.

### Construction of Metabolites Risk Score

The Metabolites Risk Score (MRS) was constructed based on metabolites’ coefficients estimated from the Lasso regression. First, 215 metabolites with unadjusted p-value < 0.2 in the main analysis of continuous, known, metabolites were identified (model 1) and carried forward. We then applied Lasso on all metabolites jointly while adjusting for all covariates used in model 1. The penalty parameter was selected by minimizing misclassification through 5-fold cross-validation. We computed sensitivity, specificity, and accuracy for the total analytic dataset and the *APOE*-ɛ4 stratified datasets. We set decision boundary to predict MCI $P_{mci}$ as the value for which $\sum_{i=1}^{n} 1\left( p_{i}>p_{mci} \right)=n_{mci}$, where $p_{i}$ is the estimated probability of MCI for person $i=1, \ldots, n$, and $n_{mci}$ is the number of individuals with MCI in the dataset. Sixty-one metabolites were consequently selected by the algorithm and formed an MRS. Finally, model performance was compared to the covariates from model 1 without the MRS. The MRS is of the form $\sum_{k=1}^{K} M_{k}\beta_{k}$ where $M_{k}$ and $\beta_{k}$ are the concentrations and the estimated effect size of the $k$th metabolite, $k=1, \ldots, K$. Here, $M_{k}$ is the metabolite concentration on the original scale, prior to rank-normalization. Rank-normalization results in all metabolites having the same standard deviation (SD) of 1. Thus, prior to rank-normalization, we recorded the SD of each of the metabolites ($SD_{1},\ldots., SD_{K}$). We applied Lasso on rank-normalized metabolite levels to obtain$, k=1, \ldots, K$, and then scaled them to have $\beta_{k}=\beta_{k}^{lasso}/SD_{k}$. These are the coefficients that should be used to construct the MRS in a new dataset. When constructing the MRS in a new dataset, we still recommend rank-normalizing the metabolites for MRS construction because the distributions of metabolites may be highly non-normal and have a high leverage point. Therefore, to construct the MRS in a new dataset, first, compute the SDs of the metabolites in the new dataset $SD_{1}^{new},\ldots, SD_{K}^{new}$, second, rank-normalize the metabolites to obtain $M_{1}^{rn},\ldots, M_{k}^{rn}$, and third, scale them to their original scale to obtain $SD_{1}^{new}M_{1}^{rn}, \ldots, {SD_{K}^{new}M}_{K}^{rn}$. The MRS in a new dataset should then be constructed by using $\sum_{k=1}^{K} {SD_{k}^{new}M}_{k}^{rn}\beta_{k}$. The MRS coefficients $\beta_{1},\ldots, \beta_{K}$ are provided in Supplementary Table 6, and a code to compute an MRS in a new dataset is provided on the GitHub repository.

Supplementary Figure 2 visualizes the association of all 61 metabolites with MCI, global cognitive change, and the change in three cognitive tests (secondary analysis) and highlights nominal associations. Some, but not all, of the metabolites reported in Table 2 (Figure 1 Step A) were selected by Lasso. This is because Lasso tends to select only one of a group of highly correlated variables (so that, out of two correlated metabolites, only one will often be selected)^3^. In addition, because Lasso performs variable selection and estimation at the same time, it does not require a second multiple testing adjustment step, and the results include metabolite with potentially low association effect size.

Supplementary Table 1: Association between FDR-adjusted significant metabolites associated with MCI or Global Cognitive Change and DSS in our dataset in order to identify a potential proxy-phenotype.

| **Metabolite** | **All Participants** | | | ***APOE*-Ɛ4 Carriers** | | | ***APOE*-Ɛ4 Non-Carriers** | | |
| --- | --- | --- | --- | --- | --- | --- | --- | --- | --- |
|  | **Effect** | **SE** | **Unadj-p-value** | **Effect** | **SE** | **Unadj p-value** | **Effect** | **SE** | **Unadj p-value** |
| Association between MCI associated metabolites and DSS | | | | | | | | | |
| MODEL 1 | | | | | | | | | |
| 3-aminoisobutyrate | 0.53 | 0.20 | 8.91E-03 | 1.02 | 0.45 | 2.17E-02 | 0.42 | 0.24 | 7.97E-02 |
| 1-arachidonoyl-GPE (20:4n6) | -0.20 | 0.30 | 4.93E-01 | 0.40 | 0.40 | 3.19E-01 | -0.35 | 0.37 | 3.53E-01 |
| Glycolithocholate sulfate | -0.27 | 0.31 | 3.80E-01 | 0.29 | 0.42 | 4.82E-01 | -0.45 | 0.39 | 2.45E-01 |
| 1-palmitoyl-2-arachidonoyl-GPE (16:0/20:4) | -0.23 | 0.30 | 4.52E-01 | 0.43 | 0.46 | 3.52E-01 | -0.36 | 0.35 | 3.05E-01 |
| Quinolinate | 0.83 | 0.29 | 3.78E-03 | 0.02 | 0.43 | 9.69E-01 | 1.07 | 0.35 | 2.21E-03 |
| 9,10-DiHOME | -0.19 | 0.29 | 5.24E-01 | 0.61 | 0.42 | 1.48E-01 | -0.48 | 0.35 | 1.73E-01 |
| Glycodeoxycholate sulfate | -0.11 | 0.31 | 7.26E-01 | 0.16 | 0.39 | 6.81E-01 | -0.24 | 0.39 | 5.35E-01 |
| 2-hydroxyoctanoate | 0.13 | 0.32 | 6.82E-01 | -0.32 | 0.38 | 4.07E-01 | 0.16 | 0.38 | 6.65E-01 |
| MODEL 2 | | | | | | | | | |
| 3-aminoisobutyrate | 0.41 | 0.23 | 6.78E-02 | 0.97 | 0.44 | 2.73E-02 | 0.23 | 0.26 | 3.88E-01 |
| 3-hydroxy-3-methylglutarate | 0.13 | 0.30 | 6.76E-01 | 0.94 | 0.39 | 1.64E-02 | -0.20 | 0.35 | 5.63E-01 |
| 1-arachidonoyl-GPE (20:4n6) | -0.20 | 0.30 | 5.01E-01 | 0.54 | 0.41 | 1.90E-01 | -0.41 | 0.37 | 2.76E-01 |
| Glycolithocholate sulfate | -0.20 | 0.32 | 5.30E-01 | 0.34 | 0.41 | 4.02E-01 | -0.40 | 0.40 | 3.19E-01 |
| palmitoleoylGPC161 | -0.19 | 0.34 | 5.78E-01 | 0.08 | 0.48 | 8.63E-01 | -0.29 | 0.40 | 4.71E-01 |
| MODEL 3 | | | | | | | | | |
| 3-aminoisobutyrate | 0.42 | 0.23 | 5.96E-02 | 1.00 | 0.43 | 2.09E-02 | 0.26 | 0.27 | 3.29E-01 |
| 3-hydroxy-3-methylglutarate | 0.14 | 0.30 | 6.35E-01 | 1.01 | 0.39 | 9.43E-03 | -0.20 | 0.34 | 5.51E-01 |
| Isocitrate | -0.52 | 0.30 | 8.05E-02 | -0.42 | 0.50 | 4.00E-01 | -0.61 | 0.35 | 7.98E-02 |
| Sulfate | 0.36 | 0.27 | 1.81E-01 | 1.17 | 0.42 | 5.86E-03 | 0.16 | 0.34 | 6.26E-01 |
| Tartarate | 0.06 | 0.23 | 7.95E-01 | 0.00 | 0.38 | 9.94E-01 | 0.04 | 0.29 | 8.82E-01 |
| 1-arachidonoyl-GPE (20:4n6) | -0.23 | 0.30 | 4.49E-01 | 0.56 | 0.41 | 1.73E-01 | -0.44 | 0.37 | 2.41E-01 |
| Glycolithocholate sulfate | -0.21 | 0.32 | 5.05E-01 | 0.35 | 0.41 | 3.88E-01 | -0.42 | 0.40 | 2.86E-01 |
| 1-palmitoleoyl-GPC (16:1) | -0.20 | 0.34 | 5.64E-01 | 0.09 | 0.49 | 8.59E-01 | -0.30 | 0.40 | 4.51E-01 |
| Association between Global Cognitive Change associated metabolites and DSS | | | | | | | | | |
| MODEL 1 | | | | | | | | | |
| Kynurenine | 0.79 | 0.29 | 5.92E-03 | -0.22 | 0.47 | 6.43E-01 | 1.04 | 0.34 | 2.00E-03 |
| 7-methylguanine | 0.95 | 0.27 | 4.57E-04 | 0.36 | 0.42 | 3.92E-01 | 1.08 | 0.31 | 5.59E-04 |
| Quinolinate | 0.83 | 0.29 | 3.78E-03 | 0.02 | 0.43 | 9.69E-01 | 1.07 | 0.35 | 2.21E-03 |
| Creatinine | 1.11 | 0.37 | 2.48E-03 | 0.35 | 0.58 | 5.44E-01 | 1.38 | 0.43 | 1.43E-03 |
| Gamma-glutamylleucine | 0.61 | 0.27 | 2.26E-02 | -0.25 | 0.46 | 5.90E-01 | 0.98 | 0.33 | 3.25E-03 |
| Gamma glutamylvaline | 0.82 | 0.33 | 1.25E-02 | 0.43 | 0.46 | 3.43E-01 | 0.95 | 0.40 | 1.68E-02 |
| N-acetylcarnosine | 1.36 | 0.34 | 6.63E-05 | 0.35 | 0.59 | 5.59E-01 | 1.68 | 0.39 | 1.72E-05 |
| Thymol sulfate | 0.91 | 0.26 | 5.31E-04 | -0.43 | 0.40 | 2.81E-01 | 1.15 | 0.30 | 1.08E-04 |

Abbreviations: MCI mild cognitive impairment, DSS digit symbol substitution test score.

Highlighting represents a significant nominal association between the metabolite and the corresponding proxy phenotype.

Supplementary Table 2: Association between FDR-adjusted significant metabolites associated with MCI or Global Cognitive Change and SEVLT in our dataset in order to identify a potential proxy-phenotype.

| **Metabolite** | **All Participants** | | | ***APOE*-Ɛ4 Carriers** | | | ***APOE*-Ɛ4 Non-Carriers** | | |
| --- | --- | --- | --- | --- | --- | --- | --- | --- | --- |
|  | **Effect** | **SE** | **Unadj-p-value** | **Effect** | **SE** | **Unadj p-value** | **Effect** | **SE** | **Unadj p-value** |
| Association between MCI metabolites and SEVLT | | | | | | | | | |
| MODEL 1 | | | | | | | | | |
| 3-aminoisobutyrate | -0.05 | 0.10 | 6.08E-01 | 0.02 | 0.17 | 9.11E-01 | -0.07 | 0.11 | 5.33E-01 |
| 1-arachidonoyl-GPE (20:4n6) | -0.04 | 0.11 | 6.73E-01 | 0.02 | 0.18 | 9.19E-01 | -0.03 | 0.13 | 7.94E-01 |
| Glycolithocholate sulfate | -0.05 | 0.09 | 6.14E-01 | -0.04 | 0.18 | 8.19E-01 | -0.06 | 0.10 | 5.48E-01 |
| 1-palmitoyl-2-arachidonoyl-GPE (16:0/20:4) | 0.11 | 0.11 | 2.90E-01 | 0.03 | 0.20 | 8.67E-01 | 0.14 | 0.12 | 2.45E-01 |
| Quinolinate | 0.14 | 0.09 | 1.23E-01 | 0.09 | 0.18 | 6.04E-01 | 0.13 | 0.10 | 2.21E-01 |
| 9,10-DiHOME | 0.05 | 0.12 | 6.63E-01 | -0.14 | 0.18 | 4.24E-01 | 0.10 | 0.14 | 4.69E-01 |
| Glycodeoxycholate sulfate | 0.06 | 0.11 | 5.89E-01 | 0.13 | 0.20 | 5.15E-01 | 0.03 | 0.12 | 7.94E-01 |
| 2-hydroxyoctanoate | 0.05 | 0.09 | 5.92E-01 | -0.15 | 0.16 | 3.59E-01 | 0.12 | 0.11 | 2.74E-01 |
| MODEL 2 | | | | | | | | | |
| 3-aminoisobutyrate | -0.04 | 0.10 | 6.60E-01 | 0.04 | 0.16 | 8.15E-01 | -0.06 | 0.11 | 5.88E-01 |
| 3-hydroxy3methylglutarate | 0.07 | 0.11 | 5.56E-01 | -0.19 | 0.18 | 3.04E-01 | 0.14 | 0.13 | 2.58E-01 |
| 1-arachidonoyl-GPE (20:4n6) | -0.05 | 0.10 | 6.14E-01 | 0.01 | 0.17 | 9.51E-01 | -0.04 | 0.13 | 7.40E-01 |
| Glycolithocholate sulfate | -0.07 | 0.09 | 4.67E-01 | -0.08 | 0.18 | 6.39E-01 | -0.09 | 0.10 | 3.79E-01 |
| 1-palmitoleoyl-GPC (16:1) | -0.02 | 0.09 | 8.05E-01 | 0.02 | 0.18 | 8.89E-01 | -0.04 | 0.11 | 7.23E-01 |
| MODEL 3 | | | | | | | | | |
| 3-aminoisobutyrate | -0.04 | 0.09 | 6.76E-01 | 0.05 | 0.16 | 7.59E-01 | -0.05 | 0.11 | 6.59E-01 |
| 3-hydroxy-3-methylglutarate | 0.08 | 0.11 | 4.86E-01 | -0.19 | 0.19 | 3.12E-01 | 0.15 | 0.13 | 2.35E-01 |
| Isocitrate | 0.05 | 0.10 | 6.57E-01 | 0.10 | 0.17 | 5.35E-01 | 0.01 | 0.12 | 9.15E-01 |
| Sulfate | 0.10 | 0.12 | 3.68E-01 | 0.33 | 0.18 | 6.67E-02 | 0.02 | 0.13 | 8.55E-01 |
| Tartarate | 0.11 | 0.10 | 2.70E-01 | -0.09 | 0.18 | 5.99E-01 | 0.15 | 0.12 | 2.25E-01 |
| 1-arachidonoyl-GPE (20:4n6) | -0.06 | 0.10 | 5.43E-01 | 0.02 | 0.16 | 8.98E-01 | -0.05 | 0.13 | 7.07E-01 |
| Glycolithocholate sulfate | -0.07 | 0.09 | 4.51E-01 | -0.06 | 0.18 | 7.41E-01 | -0.09 | 0.10 | 3.49E-01 |
| 1-palmitoleoyl-GPC (16:1) | -0.02 | 0.09 | 7.88E-01 | 0.06 | 0.17 | 7.34E-01 | -0.04 | 0.11 | 6.90E-01 |
| Association between Global Cognitive Change metabolites and SEVLT | | | | | | | | | |
| MODEL 1 | | | | | | | | | |
| Kynurenine | 0.26 | 0.09 | 2.51E-03 | -0.01 | 0.17 | 9.33E-01 | 0.30 | 0.10 | 2.92E-03 |
| 7-methylguanine | 0.05 | 0.11 | 6.73E-01 | -0.27 | 0.17 | 1.22E-01 | 0.12 | 0.12 | 2.95E-01 |
| Quinolinate | 0.14 | 0.09 | 1.23E-01 | 0.09 | 0.18 | 6.04E-01 | 0.13 | 0.10 | 2.21E-01 |
| Creatinine | -0.02 | 0.11 | 8.59E-01 | -0.13 | 0.20 | 5.12E-01 | 0.02 | 0.13 | 8.68E-01 |
| Gamma-glutamylleucine | 0.16 | 0.12 | 2.03E-01 | 0.07 | 0.19 | 7.22E-01 | 0.16 | 0.14 | 2.61E-01 |
| Gamma-glutamylvaline | 0.16 | 0.09 | 8.11E-02 | 0.29 | 0.14 | 3.90E-02 | 0.12 | 0.11 | 2.96E-01 |
| N-acetylcarnosine | 0.05 | 0.14 | 7.36E-01 | 0.25 | 0.22 | 2.53E-01 | 0.01 | 0.17 | 9.53E-01 |
| Thymol sulfate | 0.06 | 0.10 | 4.98E-01 | 0.20 | 0.18 | 2.47E-01 | 0.04 | 0.11 | 6.95E-01 |

Abbreviations: MCI mild cognitive impairment, SEVLT Spanish English Verbal Learning Test.

Highlighting represents a significant nominal association between the metabolite and the corresponding proxy phenotype.

Supplementary Table 3: Association between FDR-adjusted significant metabolites associated with MCI or Global Cognitive Change and Word Fluency in our dataset in order to identify a potential proxy-phenotype.

| **Metabolite** | **All Participants** | | | ***APOE*-Ɛ4 Carriers** | | | ***APOE*-Ɛ4 Non-Carriers** | | |
| --- | --- | --- | --- | --- | --- | --- | --- | --- | --- |
|  | **Effect** | **SE** | **Unadj-p-value** | **Effect** | **SE** | **Unadj p-value** | **Effect** | **SE** | **Unadj p-value** |
| Association between MCI metabolites and WF | | | | | | | | | |
| MODEL 1 | | | | | | | | | |
| 3-aminoisobutyrate | 0.13 | 0.22 | 5.60E-01 | 1.22 | 0.44 | 5.16E-03 | -0.12 | 0.23 | 5.97E-01 |
| 1-arachidonoyl-GPE (20:4n6) | -0.29 | 0.23 | 2.06E-01 | 0.40 | 0.38 | 2.93E-01 | -0.43 | 0.27 | 1.03E-01 |
| Glycolithocholate sulfate | -0.27 | 0.21 | 2.00E-01 | 0.60 | 0.49 | 2.21E-01 | -0.54 | 0.23 | 2.17E-02 |
| 1-palmitoyl-2-arachidonoyl-GPE (16:0/20:4) | -0.19 | 0.21 | 3.59E-01 | 0.22 | 0.47 | 6.31E-01 | -0.26 | 0.24 | 2.87E-01 |
| Quinolinate | 0.37 | 0.19 | 4.65E-02 | 0.19 | 0.45 | 6.76E-01 | 0.42 | 0.20 | 3.63E-02 |
| 9,10-DiHOME | -0.03 | 0.22 | 9.02E-01 | 0.44 | 0.57 | 4.42E-01 | -0.19 | 0.22 | 3.99E-01 |
| Glycodeoxycholate sulfate | -0.16 | 0.18 | 3.86E-01 | 0.41 | 0.41 | 3.09E-01 | -0.36 | 0.21 | 8.22E-02 |
| 2-hydroxyoctanoate | 0.21 | 0.21 | 3.18E-01 | -0.07 | 0.50 | 8.91E-01 | 0.27 | 0.21 | 1.89E-01 |
| MODEL 2 | | | | | | | | | |
| 3-aminoisobutyrate | 0.12 | 0.22 | 5.82E-01 | 1.22 | 0.45 | 6.22E-03 | -0.18 | 0.24 | 4.61E-01 |
| 3-hydroxy3methylglutarate | -0.02 | 0.21 | 9.43E-01 | -0.10 | 0.53 | 8.44E-01 | 0.01 | 0.23 | 9.55E-01 |
| 1-arachidonoyl-GPE (20:4n6) | -0.29 | 0.23 | 2.10E-01 | 0.51 | 0.40 | 2.00E-01 | -0.45 | 0.26 | 8.27E-02 |
| Glycolithocholate sulfate | -0.30 | 0.21 | 1.43E-01 | 0.60 | 0.50 | 2.32E-01 | -0.60 | 0.22 | 7.37E-03 |
| 1-palmitoleoyl-GPC (16:1) | -0.35 | 0.21 | 9.52E-02 | -0.53 | 0.43 | 2.18E-01 | -0.28 | 0.25 | 2.57E-01 |
| MODEL 3 | | | | | | | | | |
| 3-aminoisobutyrate | 0.12 | 0.20 | 5.62E-01 | 1.26 | 0.43 | 3.30E-03 | -0.16 | 0.22 | 4.71E-01 |
| 3-hydroxy-3-methylglutarate | 0.01 | 0.21 | 9.60E-01 | 0.00 | 0.52 | 9.94E-01 | 0.01 | 0.22 | 9.67E-01 |
| Isocitrate | 0.21 | 0.21 | 3.37E-01 | -0.21 | 0.53 | 6.86E-01 | 0.21 | 0.22 | 3.30E-01 |
| Sulfate | 0.19 | 0.24 | 4.19E-01 | 0.66 | 0.45 | 1.38E-01 | 0.02 | 0.27 | 9.41E-01 |
| Tartarate | -0.10 | 0.24 | 6.87E-01 | -0.80 | 0.53 | 1.31E-01 | 0.16 | 0.26 | 5.34E-01 |
| 1-arachidonoyl-GPE (20:4n6) | -0.32 | 0.23 | 1.60E-01 | 0.38 | 0.40 | 3.41E-01 | -0.48 | 0.26 | 6.09E-02 |
| Glycolithocholate sulfate | -0.35 | 0.20 | 7.87E-02 | 0.61 | 0.49 | 2.10E-01 | -0.65 | 0.21 | 2.24E-03 |
| 1-palmitoleoyl-GPC (16:1) | -0.37 | 0.21 | 7.25E-02 | -0.67 | 0.42 | 1.12E-01 | -0.29 | 0.24 | 2.17E-01 |
| Association between Global Cognitive Change metabolites and WF | | | | | | | | | |
| MODEL 1 | | | | | | | | | |
| Kynurenine | 0.39 | 0.20 | 5.27E-02 | 0.20 | 0.44 | 6.47E-01 | 0.40 | 0.22 | 6.77E-02 |
| 7-methylguanine | 0.29 | 0.18 | 9.70E-02 | 0.16 | 0.34 | 6.51E-01 | 0.37 | 0.20 | 6.23E-02 |
| Quinolinate | 0.37 | 0.19 | 4.65E-02 | 0.19 | 0.45 | 6.76E-01 | 0.42 | 0.20 | 3.63E-02 |
| Creatinine | 0.16 | 0.21 | 4.47E-01 | 0.01 | 0.52 | 9.81E-01 | 0.28 | 0.22 | 2.11E-01 |
| Gamma-glutamylleucine | 0.32 | 0.23 | 1.63E-01 | 0.14 | 0.52 | 7.86E-01 | 0.38 | 0.26 | 1.45E-01 |
| Gamma-glutamylvaline | 0.38 | 0.21 | 6.50E-02 | 0.03 | 0.51 | 9.50E-01 | 0.48 | 0.22 | 3.20E-02 |
| N-acetylcarnosine | 0.24 | 0.24 | 3.16E-01 | 0.42 | 0.59 | 4.84E-01 | 0.25 | 0.25 | 3.13E-01 |
| Thymol sulfate | -0.20 | 0.19 | 2.82E-01 | -0.74 | 0.51 | 1.49E-01 | -0.08 | 0.20 | 6.74E-01 |

Abbreviations: MCI mild cognitive impairment, WF word fluency test.

Highlighting represents a significant nominal association between the metabolite and the corresponding proxy phenotype.

Supplementary Table 4: Association between FDR-adjusted significant metabolites (previously associated with MCI or Global Cognitive Change) with a chosen proxy-phenotype in our analytic dataset, within the total population and stratified by *APOE*-Ɛ4 carrier status.

| **Metabolite** | **Model** | **All Participants** | | | | | ***APOE*-Ɛ4 Carriers** | | | | |  | ***APOE*-Ɛ4 Non-Carriers** | | | |
| --- | --- | --- | --- | --- | --- | --- | --- | --- | --- | --- | --- | --- | --- | --- | --- | --- |
|  |  | **Effect** | **SE** | **Unadj-p-value** | **FDR adj p-value** | **Proxy Outcome** | **Effect** | **SE** | **Unadj-p-value** | **FDR adj p-value** | **Proxy Outcome** | **Effect** | **SE** | **Unadj-p-value** | **FDR adj p-value** | **Proxy Outcome** |
| **Metabolites associated with MCI** | | | | | | | | | | | | | | | | |
| 3-aminoisobutyrate | Model 1 | -0.04 | 0.01 | 1.26E-02 | 1.00E+00 | NA | -0.69 | 0.17 | 3.00E-05 | 1.76E-02 | WF | -0.21 | 0.10 | 4.19E-02 | 1.00E+00 | NA |
| 1-arachidonoyl-GPE (20:4n6) | Model 1 | 0.03 | 0.01 | 1.44E-02 | 1.00E+00 | NA | -0.20 | 0.15 | 1.80E-01 | 1.00E+00 | NA | 0.40 | 0.10 | 4.00E-05 | 1.71E-02 | NA |
| Glycolithocholate sulfate | Model 1 | 0.04 | 0.01 | 1.12E-03 | 1.00E+00 | NA | -0.29 | 0.20 | 1.38E-01 | 1.00E+00 | NA | 0.42 | 0.11 | 5.00E-05 | 1.71E-02 | WF |
| 1-palmitoyl-2-arachidonoyl-GPE (16:0/20:4) | Model 1 | 0.03 | 0.01 | 6.63E-03 | 1.00E+00 | NA | -0.04 | 0.16 | 7.82E-01 | 1.00E+00 | NA | 0.40 | 0.11 | 2.20E-04 | 3.52E-02 | NA |
| Quinolinate | Model 1 | -0.03 | 0.01 | 6.66E-04 | 5.99E-01 | NA | -0.13 | 0.17 | 4.37E-01 | 1.00E+00 | NA | -0.33 | 0.09 | 4.30E-04 | 4.62E-02 | DSS |
| 9,10-DiHOME | Model 1 | -0.04 | 0.01 | 4.16E-04 | 3.74E-01 | NA | -0.13 | 0.20 | 5.23E-01 | 1.00E+00 | NA | -0.38 | 0.11 | 5.10E-04 | 4.66E-02 | NA |
| Glycodeoxycholate sulfate | Model 1 | 0.04 | 0.01 | 8.64E-04 | 7.77E-01 | NA | 0.02 | 0.20 | 9.23E-01 | 1.00E+00 | NA | 0.34 | 0.09 | 1.70E-04 | 3.52E-02 | NA |
| 2-hydroxyoctanoate | Model 1 | -0.04 | 0.01 | 2.06E-04 | 1.86E-01 | NA | -0.08 | 0.14 | 5.64E-01 | 1.00E+00 | NA | -0.36 | 0.10 | 3.10E-04 | 3.93E-02 | NA |
| 3-aminoisobutyrate | Model 2 | -0.04 | 0.02 | 1.93E-02 | 1.00E+00 | NA | -0.66 | 0.14 | 1.90E-01 | 8.50E-04 | WF | -0.17 | 0.11 | 1.11E-01 | 1.00E+00 | NA |
| 3-hydroxy-3-methylglutarate | Model 2 | -0.02 | 0.01 | 9.42E-02 | 1.00E+00 | NA | -0.68 | 0.16 | 1.00E-05 | 4.27E-03 | DSS | -0.06 | 0.11 | 6.16E-01 | 1.00E+00 | NA |
| 1-arachidonoyl-GPE (20:4n6) | Model 2 | 0.03 | 0.01 | 3.92E-02 | 1.00E+00 | NA | -0.19 | 0.16 | 2.16E-01 | 1.00E+00 | NA | 0.42 | 0.10 | 4.00E-05 | 2.69E-02 | NA |
| Glycolithocholate sulfate | Model 2 | 0.04 | 0.01 | 2.60E-03 | 1.00E+00 | NA | -0.27 | 0.18 | 1.46E-01 | 1.00E+00 | NA | 0.39 | 0.11 | 2.20E-04 | 4.78E-02 | WF |
| 1-palmitoleoyl-GPC (16:1) | Model 2 | 0.03 | 0.02 | 4.40E-02 | 1.00E+00 | NA | -0.20 | 0.13 | 1.14E-01 | 1.00E+00 | NA | 0.42 | 0.11 | 1.60E-04 | 4.78E-02 | NA |
| 3-aminoisobutyrate | Model 3 | -0.04 | 0.02 | 1.38E-02 | 1.00E+00 | NA | -0.68 | 0.16 | 1.00E-05 | 4.45E-03 | WF | -0.19 | 0.11 | 1.00E-01 | 1.00E+00 | NA |
| 3-hydroxy-3-methylglutarate | Model 3 | -0.02 | 0.01 | 9.53E-02 | 1.00E+00 | NA | -0.73 | 0.17 | 1.00E-05 | 4.45E-03 | DSS | -0.05 | 0.11 | 6.78E-01 | 1.00E+00 | NA |
| Isocitrate | Model 3 | 0.01 | 0.01 | 5.44E-01 | 1.00E+00 | NA | 0.57 | 0.16 | 2.60E-04 | 3.38E-02 | NA | 0.04 | 0.12 | 7.49E-01 | 1.00E+00 | NA |
| Sulfate | Model 3 | -0.01 | 0.01 | 4.19E-01 | 1.00E+00 | NA | -0.52 | 0.14 | 1.70E-04 | 2.72E-02 | DSS | 0.08 | 0.12 | 4.82E-01 | 1.00E+00 | NA |
| Tartarate | Model 3 | 0.00 | 0.01 | 7.72E-01 | 1.00E+00 | NA | 0.52 | 0.14 | 1.70E-04 | 2.72E-02 | NA | -0.14 | 0.10 | 1.64E-01 | 1.00E+00 | NA |
| 1-arachidonoyl-GPE (20:4n6) | Model 3 | 0.03 | 0.01 | 4.29E-02 | 1.00E+00 | NA | -0.22 | 0.16 | 1.65E-01 | 1.00E+00 | NA | 0.42 | 0.10 | 5.00E-05 | 2.01E-02 | NA |
| Glycolithocholate sulfate | Model 3 | 0.04 | 0.01 | 3.01E-03 | 1.00E+00 | NA | -0.29 | 0.18 | 1.09E-01 | 1.00E+00 | NA | 0.39 | 0.10 | 1.50E-04 | 3.15E-02 | WF |
| 1-palmitoleoyl-GPC (16:1) | Model 3 | 0.03 | 0.02 | 4.29E-02 | 1.00E+00 | NA | -0.18 | 0.13 | 1.63E-01 | 1.00E+00 | NA | 0.42 | 0.10 | 6.00E-05 | 2.01E-02 | NA |
| **Metabolites associated with Global Cognitive Change** | | | | | | | | | | | | | | | | |
| Kynurenine | Model 1 | 0.14 | 0.03 | 4.00E-05 | 2.43E-02 | SEVLT | 0.04 | 0.06 | 5.24E-01 | 1.00E+00 | NA | 0.16 | 0.04 | 6.00E-05 | 1.22E-02 | DSS |
| 7-methylguanine | Model 1 | 0.12 | 0.03 | 2.10E-04 | 4.45E-02 | DSS | 0.03 | 0.06 | 6.35E-01 | 1.00E+00 | NA | 0.15 | 0.04 | 3.00E-05 | 1.22E-02 | DSS |
| Quinolinate | Model 1 | 0.13 | 0.03 | 8.00E-05 | 2.43E-02 | DSS | 0.06 | 0.06 | 3.17E-01 | 1.00E+00 | NA | 0.15 | 0.04 | 1.20E-04 | 1.86E-02 | DSS |
| Creatinine | Model 1 | 0.12 | 0.04 | 3.46E-03 | 1.00E+00 | NA | 0.02 | 0.07 | 7.99E-01 | 1.00E+00 | NA | 0.16 | 0.05 | 4.20E-04 | 4.45E-02 | DSS |
| Gamma-glutamylleucine | Model 1 | 0.11 | 0.03 | 6.60E-04 | 5.93E-01 | NA | -0.02 | 0.07 | 7.51E-01 | 1.00E+00 | NA | 0.16 | 0.04 | 4.00E-05 | 1.22E-02 | DSS |
| Gamma-glutamylvaline | Model 1 | 0.13 | 0.04 | 3.53E-04 | 3.17E-01 | NA | 0.06 | 0.06 | 3.71E-01 | 1.00E+00 | NA | 0.15 | 0.04 | 4.90E-04 | 4.45E-02 | DSS |
| N-acetylcarnosine | Model 1 | 0.16 | 0.05 | 4.95E-04 | 4.45E-01 | NA | 0.09 | 0.08 | 2.60E-01 | 1.00E+00 | NA | 0.19 | 0.05 | 2.90E-04 | 3.68E-02 | DSS |
| Thymol sulfate | Model 1 | 0.08 | 0.03 | 6.06E-03 | 1.00E+00 | NA | -0.06 | 0.06 | 3.18E-01 | 1.00E+00 | NA | 0.11 | 0.03 | 5.90E-04 | 4.72E-02 | DSS |

Abbreviations: MCI mild cognitive impairment, DSS digit symbol substitution test score, WF word fluency test score, SEVLT Spanish English Verbal Learning Test

Highlighting represents a significant nominal association between the metabolite and the corresponding proxy phenotype.

NA indicates that no significant association was found with a proxy phenotype, thus no generalization was performed in ARIC Cohort.

Supplementary Table 5: Generalization of the FDR-adjusted significant metabolites associated with MCI or Global Cognitive Change with proxy neurocognitive outcomes in the ARIC Cohort, stratified by ancestry.

| Metabolite | Model | *APOE*-Ɛ4 Carriers | | | | *APOE*-Ɛ4 Non-Carriers | | | |
| --- | --- | --- | --- | --- | --- | --- | --- | --- | --- |
|  |  | Effect | SE | p-value | Proxy Outcome | Effect | SE | p-value | Proxy Outcome |
| **EUROPEAN AMERICAN** | | | | | | | | | |
| Metabolites associated with MCI | | | | | | | | | |
| Glycolithocholate sulfate | Model 1 |  |  |  |  | 0.11 | 0.28 | 0.71 | WF |
| Quinolinate | Model 1 |  |  |  |  | -0.19 | 0.23 | 0.41 | DSS |
| 3-hydroxy-3-methylglutarate | Model 2 | -0.06 | 0.36 | 0.87 | DSS |  |  |  |  |
| Glycolithocholate sulfate | Model 2 |  |  |  |  | 0.09 | 0.28 | 0.75 | WF |
| 3-hydroxy-3-methylglutarate | Model 3 | -0.06 | 0.36 | 0.88 | DSS |  |  |  |  |
| Sulfate | Model 3 | -0.03 | 0.36 | 0.93 | DSS |  |  |  |  |
| Glycolithocholate sulfate | Model 3 |  |  |  |  | 0.09 | 0.28 | 0.74 | WF |
| Metabolites associated with Global Cognitive Change | | | | | | | | | |
| Kynurenine | Model 1 | 0.04 | 0.05 | 0.42 | DWRT | -0.01 | 0.23 | 0.98 | DSS |
| 7-methylguanine | Model 1 | 0.03 | 0.19 | 0.87 | DSS | 0.19 | 0.23 | 0.41 | DSS |
| Quinolinate | Model 1 | -0.37 | 0.19 | 0.05 | DSS | -0.19 | 0.23 | 0.41 | DSS |
| Creatinine | Model 1 |  |  |  |  | 0.10 | 0.23 | 0.66 | DSS |
| Gamma-glutamylleucine | Model 1 |  |  |  |  | 0.33 | 0.22 | 0.13 | DSS |
| Gamma-glutamylvaline | Model 1 |  |  |  |  | 0.35 | 0.22 | 0.11 | DSS |
| N-acetylcarnosine | Model 1 |  |  |  |  | 0.30 | 0.28 | 0.30 | DSS |
| Thymol sulfate | Model 1 |  |  |  |  | 0.26 | 0.23 | 0.25 | DSS |
| **AFRICAN AMERICAN** | | | | | | | | | |
| Metabolites associated with MCI | | | | | | | | | |
| Glycolithocholate sulfate | Model 1 |  |  |  |  | -0.14 | 0.61 | 0.81 | WF |
| Quinolinate | Model 1 |  |  |  |  | -0.79 | 0.60 | 0.19 | DSS |
| 3-hydroxy-3-methylglutarate | Model 2 | -0.37 | 0.72 | 0.61 | DSS |  |  |  |  |
| Glycolithocholate sulfate | Model 2 |  |  |  |  | 0.23 | 0.62 | 0.72 | WF |
| 3-hydroxy-3-methylglutarate | Model 3 | -0.38 | 0.73 | 0.61 | DSS |  |  |  |  |
| Sulfate | Model 3 | -0.28 | 0.80 | 0.73 | DSS |  |  |  |  |
| Glycolithocholate sulfate | Model 3 |  |  |  |  | 0.17 | 0.62 | 0.79 | WF |
| Metabolites associated with Global Cognitive Change | | | | | | | | | |
| Kynurenine | Model 1 | -0.13 | 0.10 | 0.19 | DWRT | -0.06 | 0.66 | 0.93 | DSS |
| 7-methylguanine | Model 1 | 1.04 | 0.47 | 0.03 | DSS | 1.22 | 0.59 | 0.04 | DSS |
| Quinolinate | Model 1 | -0.34 | 0.48 | 0.47 | DSS | -0.79 | 0.60 | 0.19 | DSS |
| Creatinine | Model 1 |  |  |  |  | 0.76 | 0.68 | 0.27 | DSS |
| Gamma-glutamylleucine | Model 1 |  |  |  |  | 0.21 | 0.50 | 0.68 | DSS |
| Gamma-glutamylvaline | Model 1 |  |  |  |  | 0.27 | 0.52 | 0.61 | DSS |
| N-acetylcarnosine | Model 1 |  |  |  |  | 1.00 | 0.70 | 0.15 | DSS |
| Thymol sulfate | Model 1 |  |  |  |  | 0.14 | 0.59 | 0.82 | DSS |

Abbreviations: DWRT Delayed Word Recall Test, DSS digit symbol substitution test score, WF word fluency test score, SEVLT Spanish English Verbal Learning Test

The negative effect of cognitive test change is risk increasing.

Highlighting represent the significant nominal association between the metabolite and the corresponding proxy phenotype in ARIC.

Supplementary Table 6: MRS metabolite Lasso coefficients.

| **Biochemical Names** | **HMDB** | **Lasso Coefficient** |
| --- | --- | --- |
| Adenosine | HMDB00050 | -0.067 |
| Alpha-ketobutyrate | HMDB00005 | 0.132 |
| 2-aminoadipate | HMDB00510 | 0.076 |
| 3-aminoisobutyrate | HMDB03911 | -0.112 |
| 2-aminooctanoate | HMDB00991 | -0.025 |
| Betaine | HMDB00043 | -0.029 |
| Carotene diol (1) |  | 0.043 |
| Carotene diol (2) |  | 0.040 |
| Cerotoylcarnitine (C26)* | HMDB06347 | 0.064 |
| Cysteine-glutathione disulfide | HMDB00656 | -0.024 |
| 12,13-DiHOME | HMDB04705 | -0.131 |
| 1-dihomo-linolenylglycerol (20:3) |  | 0.039 |
| 2,3-dihydroxy-2-methylbutyrate | HMDB29576 | 0.030 |
| 1,2-dilinoleoyl-GPC (18:2/18:2) | HMDB08138 | -0.081 |
| Dimethylarginine (SDMA + ADMA) | HMDB01539 | -0.024 |
| Dimethylglycine | HMDB00092 | -0.087 |
| Docosadioate |  | -0.017 |
| Eicosanodioate |  | -0.129 |
| Gamma-glutamylhistidine |  | -0.123 |
| Glutamate, gamma-methyl ester | HMDB61715 | -0.051 |
| Glutarylcarnitine (C5-DC) | HMDB13130 | -0.100 |
| Glycochenodeoxycholate sulfate |  | 0.223 |
| Glycodeoxycholate | HMDB00631 | 0.048 |
| Glycosyl ceramide (d18:1/20:0, d16:1/22:0)* |  | -0.044 |
| Glycosyl-N-(2-hydroxynervonoyl)-sphingosine (d18:1/24:1(2OH))* |  | 0.009 |
| Guanidinoacetate | HMDB00128 | 0.148 |
| Hexadecanedioate | HMDB00672 | 0.064 |
| 2-hydroxyoctanoate | HMDB02264 | -0.024 |
| 3-hydroxypyridine sulfate |  | 0.024 |
| Hyocholate | HMDB00760 | -0.050 |
| Isobutyrylglycine | HMDB00730 | 0.094 |
| Lactosyl-N-palmitoyl-sphingosine (d18:1/16:0) |  | -0.059 |
| Linoleoyl-linoleoyl-glycerol (18:2/18:2) [1]* | HMDB07248 | -0.058 |
| Maleate | HMDB00176 | 0.036 |
| Methionine | HMDB00696 | -0.023 |
| N('1)-acetylspermidine | HMDB01276 | -0.010 |
| N1-methylinosine | HMDB02721 | -0.011 |
| N6,N6,N6-trimethyllysine | HMDB01325 | -0.064 |
| N6-succinyladenosine | HMDB00912 | 0.075 |
| N-acetylglucosaminylasparagine | HMDB00489 | -0.062 |
| N-trimethyl 5-aminovalerate |  | 0.070 |
| Oleoyl ethanolamide | HMDB02088 | 0.020 |
| 1-oleoyl-GPE (18:1) | HMDB11506 | 0.061 |
| 2'-O-methyluridine |  | 0.130 |
| Orotate | HMDB00226 | -0.035 |
| 5-oxoproline | HMDB00267 | -0.014 |
| 1-palmitoleoyl-GPC (16:1)* | HMDB10383 | 0.035 |
| Phenylacetylcarnitine |  | 0.050 |
| Phenylalanylglycine | HMDB28995 | -0.017 |
| Picolinate | HMDB02243 | 0.039 |
| Pimeloylcarnitine/3-methyladipoylcarnitine (C7-DC) |  | -0.026 |
| Pyroglutamine* |  | -0.001 |
| Quinolinate | HMDB00232 | -0.189 |
| Sphingomyelin (d18:1/25:0, d19:0/24:1, d20:1/23:0, d19:1/24:0)* |  | -0.117 |
| Stachydrine | HMDB04827 | 0.033 |
| 1-stearoyl-2-oleoyl-GPS (18:0/18:1) | HMDB10163 | 0.146 |
| Suberate (octanedioate) | HMDB00893 | 0.082 |
| Taurolithocholate 3-sulfate | HMDB02580 | 0.003 |
| Trigonelline (N'-methylnicotinate) | HMDB00875 | 0.033 |
| Tryptophan betaine | HMDB61115 | -0.141 |
| Ursodeoxycholate | HMDB00946 | -0.141 |

Abbreviations: MRS Metabolite risk score, HMDB Human Metabolome Database.

*Metabolite compounds that have not been officially confirmed based on a standard, but Metabolon Inc. is confident in its identity.
